# Potential Biases in Test-Negative Design Studies of COVID-19 Vaccine Effectiveness Arising from the Inclusion of Asymptomatic Individuals

**DOI:** 10.1101/2023.11.16.23298633

**Authors:** Edgar Ortiz-Brizuela, Mabel Carabali, Cong Jiang, Joanna Merckx, Denis Talbot, Mireille E. Schnitzer

## Abstract

The test-negative design (TND) is a popular method for evaluating vaccine effectiveness (VE). A “classical” TND study includes symptomatic individuals tested for the disease targeted by the vaccine to estimate VE against symptomatic infection. However, recent applications of the TND have attempted to estimate VE against infection by including all tested individuals, regardless of their symptoms. In this article, we use directed acyclic graphs and simulations to investigate potential biases in TND studies of COVID-19 VE arising from the use of this “alternative” approach, particularly when applied during periods of widespread testing. We show that the inclusion of asymptomatic individuals can potentially lead to collider stratification bias, uncontrolled confounding by health and healthcare-seeking behaviors (HSBs), and differential outcome misclassification. While our focus is on the COVID-19 setting, the issues discussed here may also be relevant in the context of other infectious diseases. This may be particularly true in scenarios where there is either a high baseline prevalence of infection, a strong correlation between HSBs and vaccination, different testing practices for vaccinated and unvaccinated individuals, or settings where both the vaccine under study attenuates symptoms of infection and diagnostic accuracy is modified by the presence of symptoms.

## Introduction

The post-licensure evaluation of COVID-19 vaccine effectiveness (VE), defined as the effect of vaccination on the risk of an infection-related outcome,(1) has provided crucial insights into questions not addressed through randomized clinical trials.(2–4) Among the study designs used to estimate VE, the test-negative design (TND) has gained popularity, in part because of its rapid and seemingly straightforward implementation.(2,5,6) In a “classical” TND study, symptomatic individuals tested for the disease targeted by the vaccine are prospectively selected, for example, from surveillance centers or hospitals.(5–8) Then, VE against symptomatic infection is estimated by comparing the odds of vaccination between patients with positive and negative test results using logistic regression.(5,7,8) However, recent literature(9–20) has also applied the term TND to studies aiming to estimate VE against infection by including all tested individuals, regardless of their symptom status (hereafter referred to as “alternative” TND) — for a clearer distinction between classical and alternative TND, see **Table 1**. Nonetheless, this approach may introduce additional threats to validity and warrants a more comprehensive evaluation.(21)

**Table 1.**
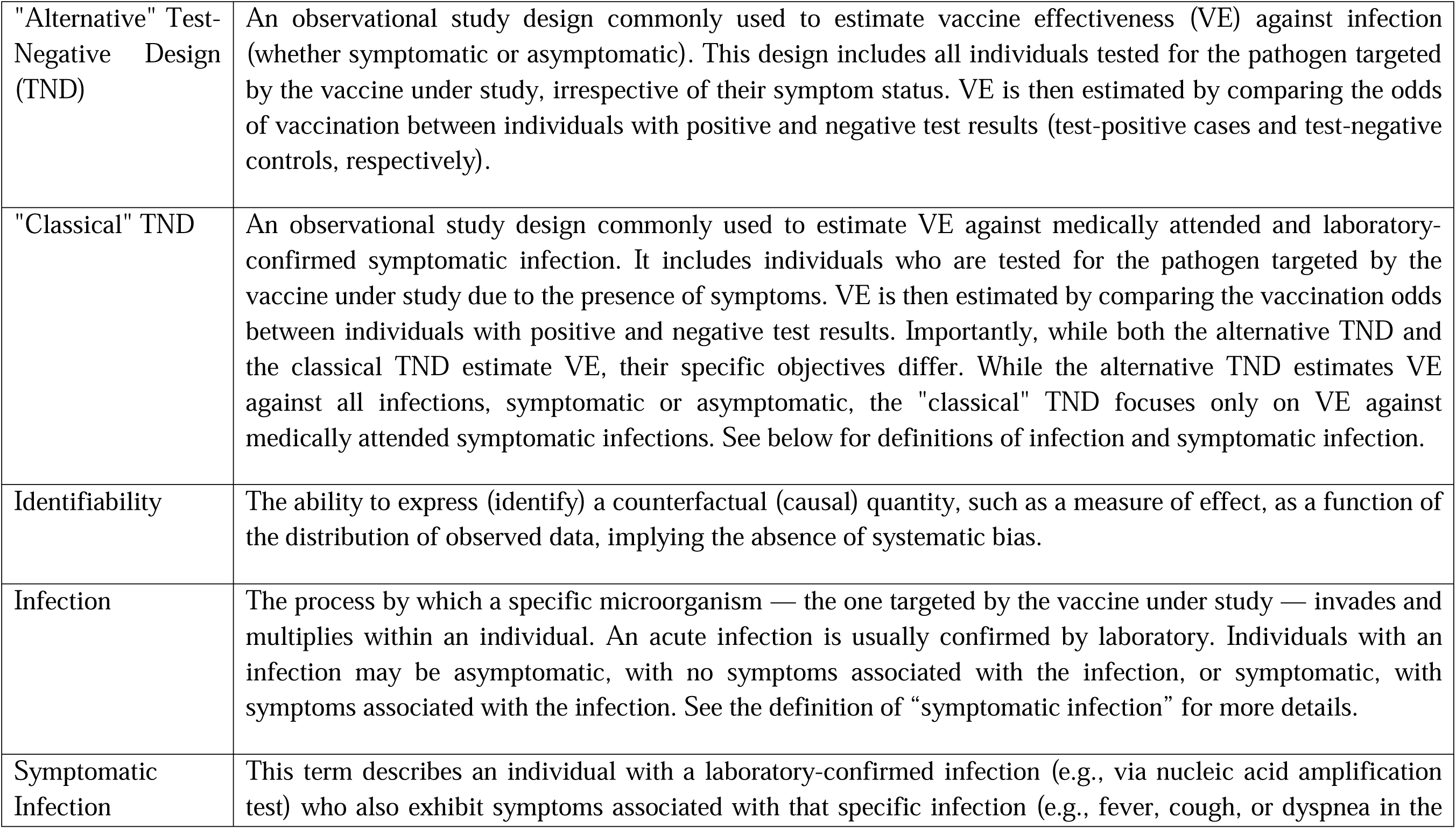
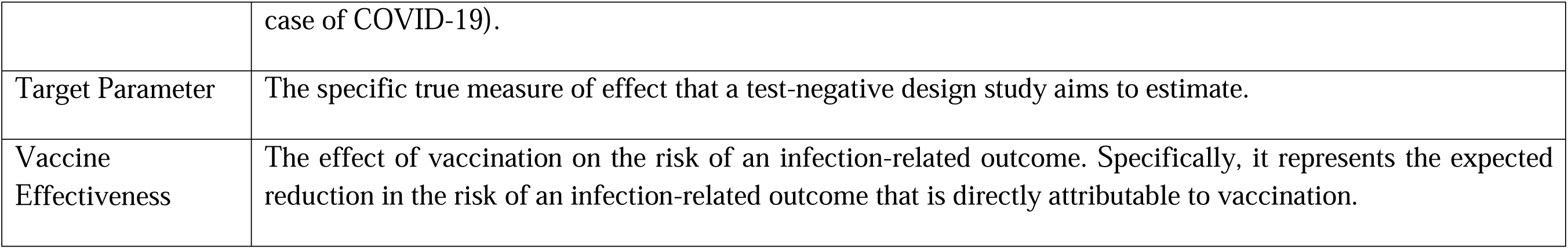
Relevant definitions used in this article.

In this article, we investigate potential biases in TND studies of COVID-19 VE arising from the use of this “alternative” approach, particularly when applied during periods of widespread testing. We begin by discussing the identifiability of two causal target parameters: 1) the risk ratio (RR) for medically attended and laboratory-confirmed symptomatic COVID-19 relative to vaccination status, referred to as *RR_COVID_* (the target parameter for the classical TND), and 2) the RR for SARS-CoV-2 infection relative to vaccination status, referred to as *RR_infect_* (the target parameter for the alternative TND) — note that throughout this manuscript we use the expression “symptomatic COVID-19” to emphasize the difference between symptomatic and asymptomatic infections. We conclude with a simulation study that aims to estimate and compare the magnitude of the bias in the odds ratio (OR) estimates for medically attended and laboratory-confirmed symptomatic COVID-19 (*OR_COVID_*) and the OR for SARS-CoV-2 infection (*OR_infect_*), relative to their respective target parameters (*RR_COVID_* and *RR_infect_*), under scenarios where a classical TND is considered valid.

## Identifiability of target parameters

In this section, we use causal directed acyclic graphs (DAGs) to examine the identifiability of the target parameters (*RR_COVID_* and *RR_infect_*).(22) By “identifiability,” we refer to the ability to express a counterfactual quantity as a function of the distribution of observed data, implying the absence of systematic biases.(23,24) However, before delving into the specifics of target parameter identifiability, and to facilitate a better understanding, we introduce the reader to a DAG based on previous work(7,8,21,25,26) that illustrates the assumed causal relationships between relevant variables in TND studies of COVID-19 VE, whether classical or alternative (**Table 2**).

**Table 2.**
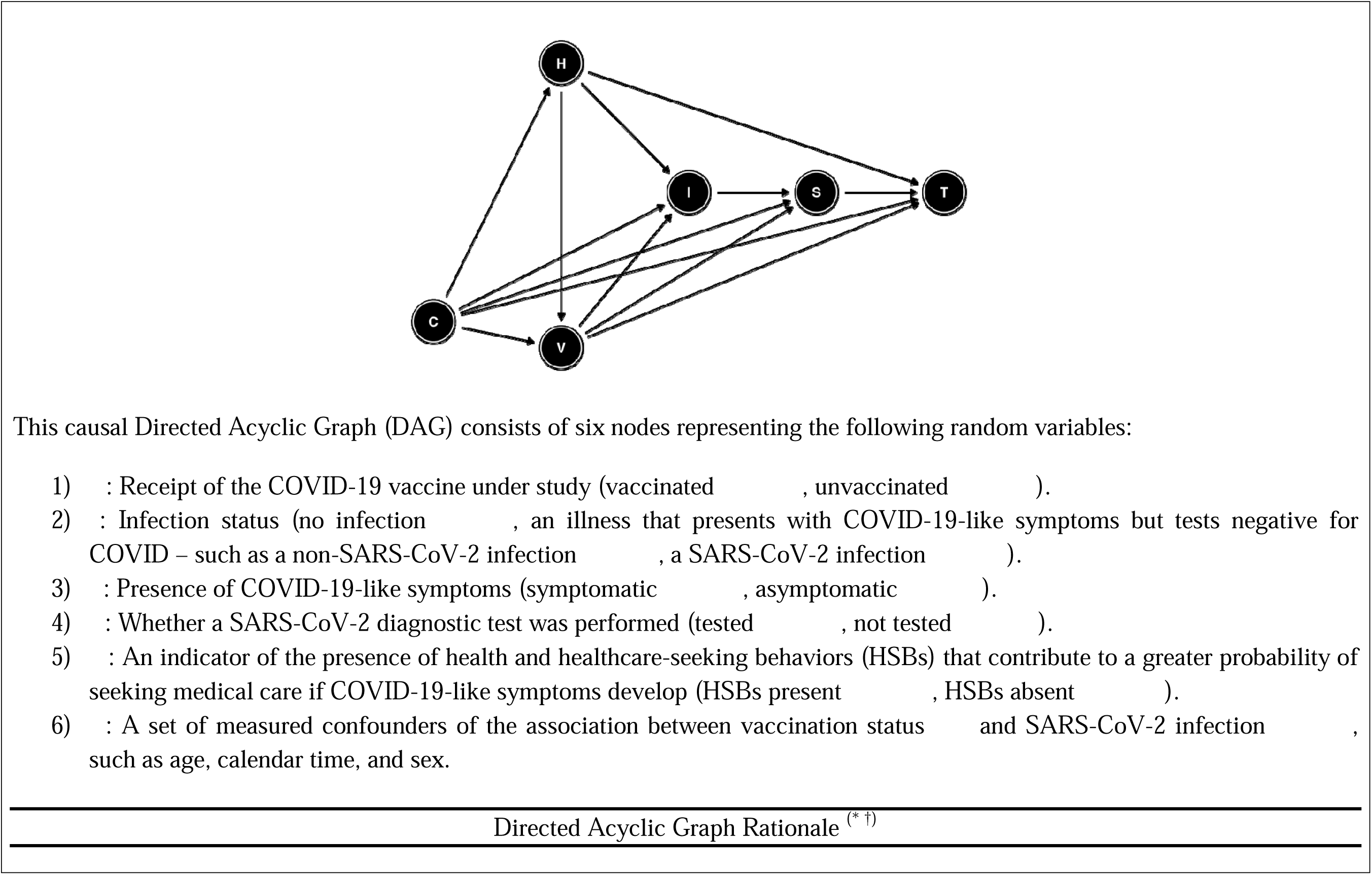
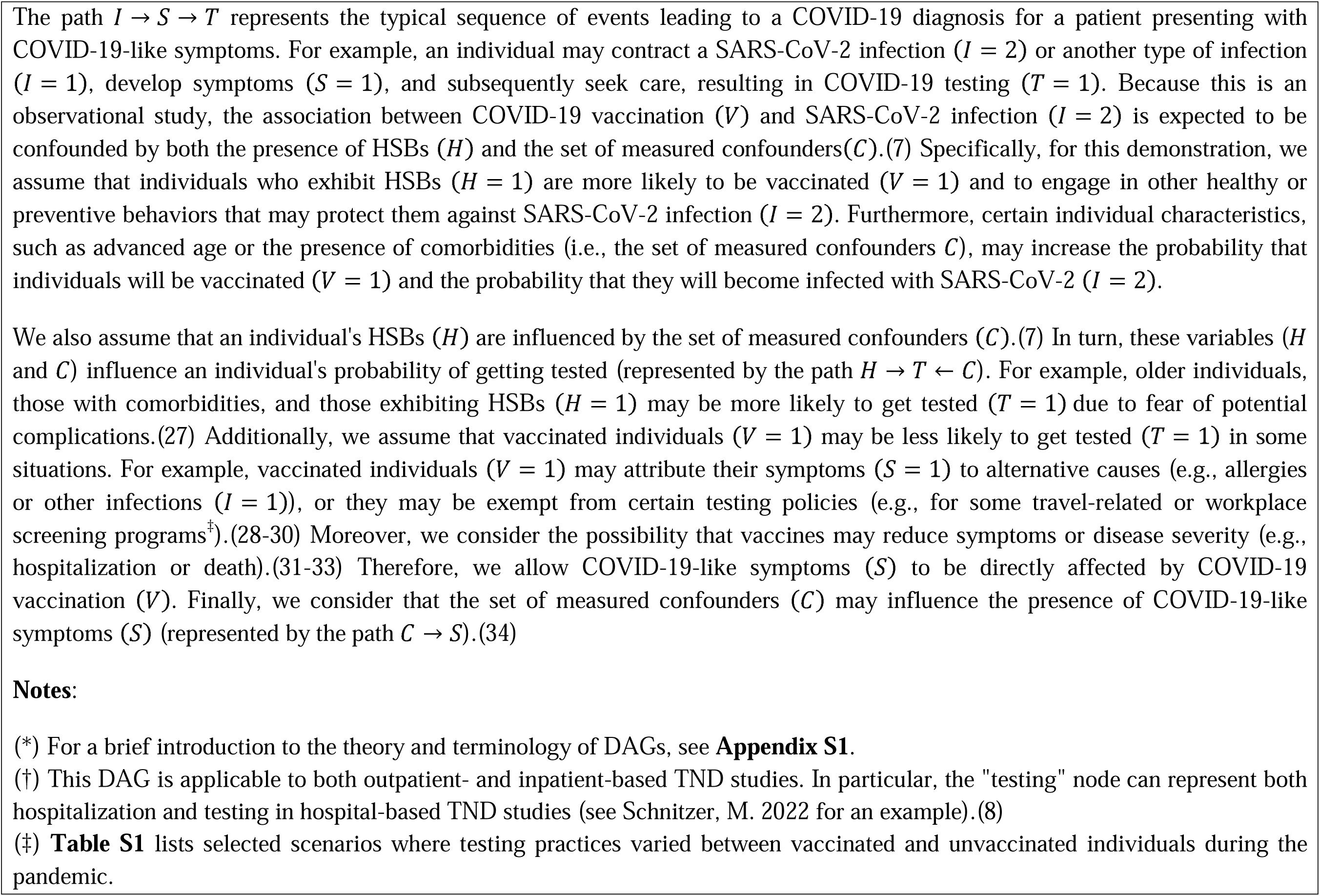
Directed acyclic graph for test-negative design studies of COVID-19 vaccine effectiveness (either classical or alternative).

### The case of the classical TND

In this context, if the test-negative state (having some illness that presents with COVID-19-like symptoms – such as an infection not targeted by the vaccine (1= 1) – and a negative test) and the vaccination status are independent conditional on covariates, the case-status OR (*OR_COVID_*) relative to vaccination status — derived from a correctly specified multivariable logistic regression model — provides an unbiased estimate of its target parameter: the conditional RR for medically attended and laboratory-confirmed symptomatic COVID-19 (1 = 2, S = 1, T = 1) relative to vaccination status (*RR_COVID_*).(8,35) Mathematical definitions for *OR_COVID_* and *RR_COVID_* are provided in **Table 3**. In hospital-based TND studies, *RR_COVID_* represents the risk ratio for medically attended infections leading to hospitalization (where T stands for “hospitalized and tested due to symptoms”).(8) On the other hand, in outpatient-based TND studies, *RR_COVID_* represents the risk ratio for medically attended infections in outpatient settings (where T stands for “accessed care and tested”).

**Table 3.**
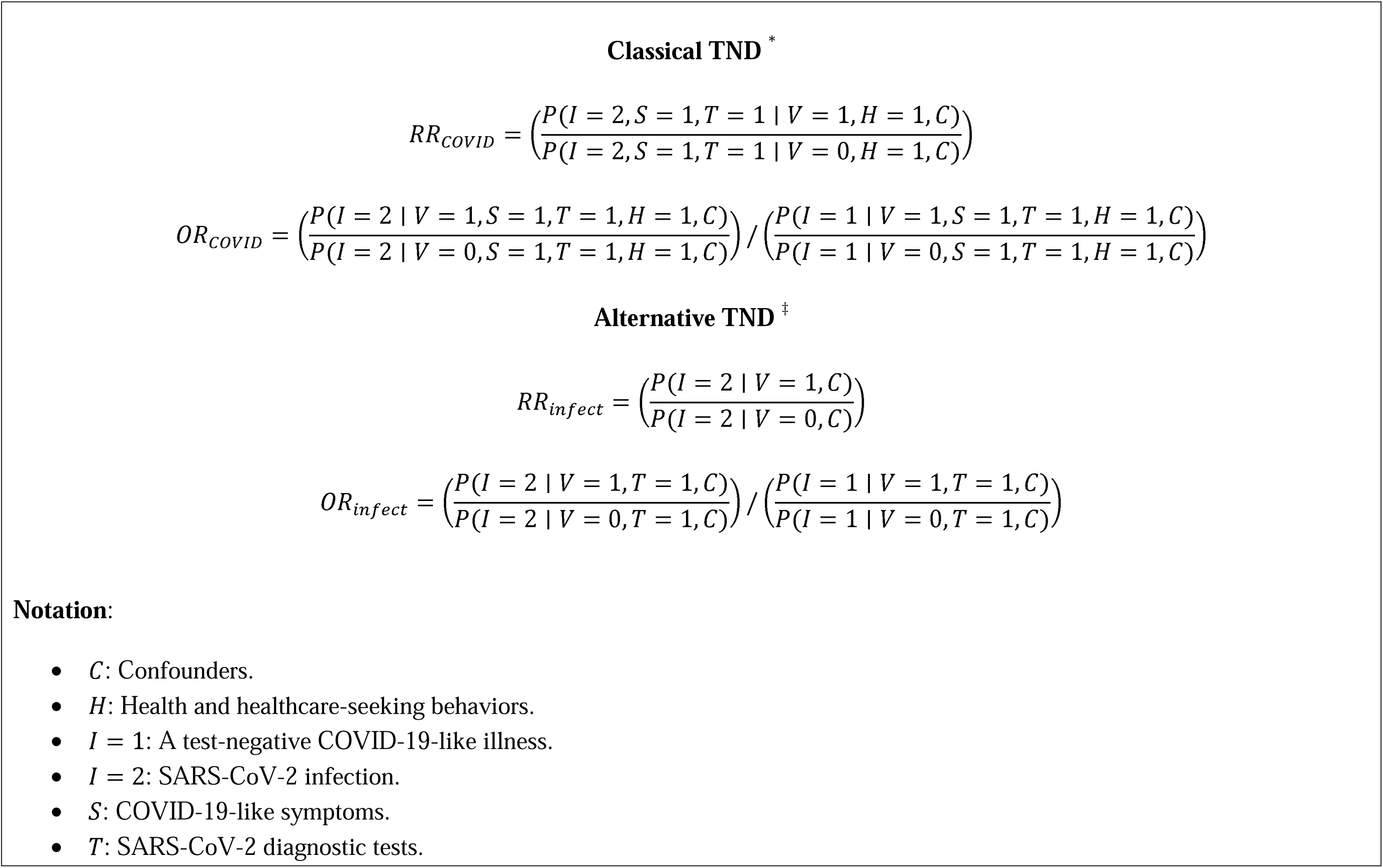
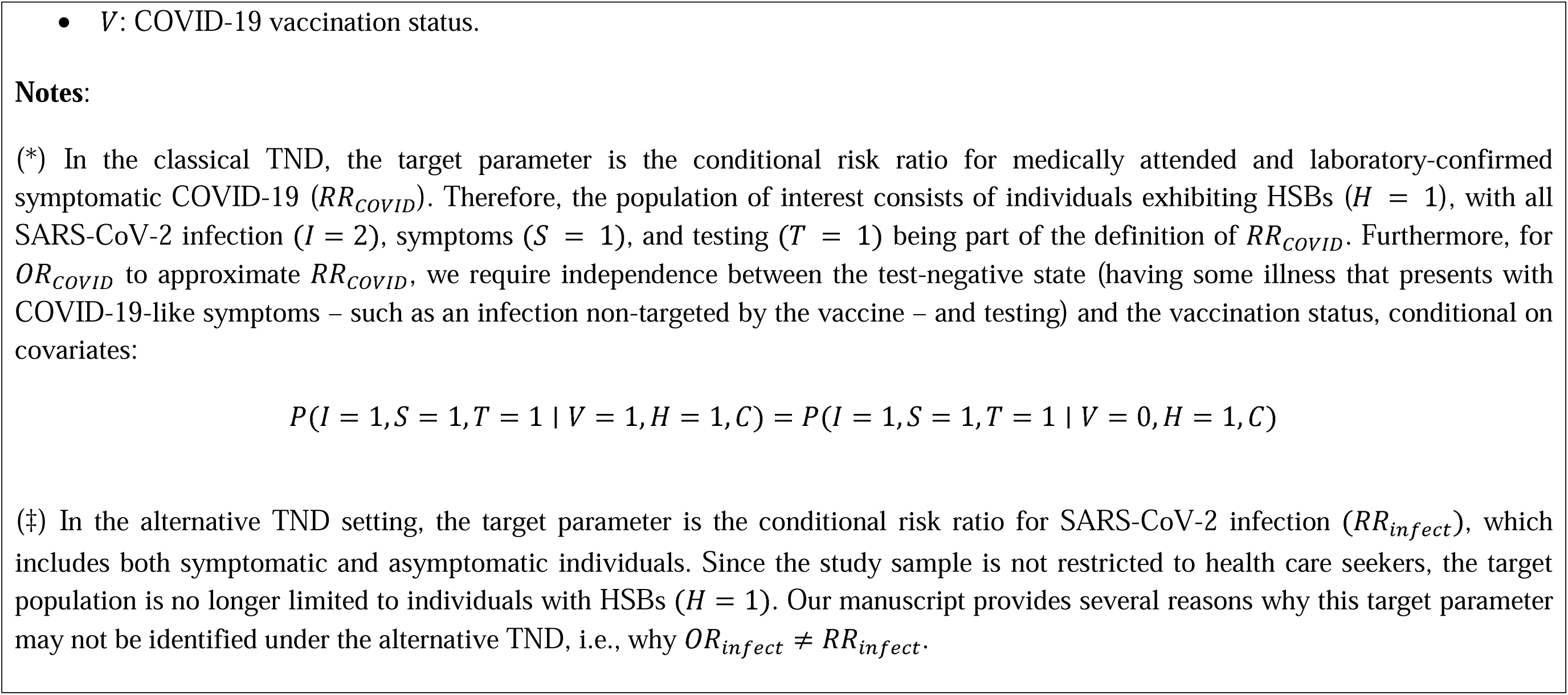
Mathematical definitions for *RR_COVID_*, *OR_COVID_*, *RR_infect_*, and *OR_infect_*.

Importantly, unlike some traditional case-control studies, the TND does not require the rare disease assumption for the OR to approximate the RR. (8) It is also noteworthy that the *OR_COVID_* estimate is often re-expressed as VE using the estimator: 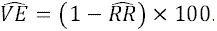. (5) However, to avoid ambiguity, throughout this manuscript and unless otherwise specified, the term VE will continue to refer to any causal effect of vaccination on the risk of an infection-related outcome, regardless of the outcome assessed.(1)

In addition, the estimands mentioned above can be given a causal interpretation if the identifiability assumptions are also met (**Appendix S2**).(8,36,37) One such assumption, known as conditional exchangeability, implies that all common causes of vaccination status (V) and outcome (medically attended and laboratory-confirmed symptomatic SARS-CoV-2 infection [1 = 2, S = 1, T = 1]) have been measured and appropriately accounted for in the analysis.(8,36–38) However, among these common causes, an individual’s health and healthcare-seeking behaviors (HSBs) are inherently unmeasured.(21) In a best-case scenario, if HSBs were deterministic (meaning that individuals who exhibit these behaviors would always seek medical care when ill, while individuals who do not exhibit HSBs would not), a classical TND study could mitigate bias related to this variable by restricting the study population to individuals who have sought medical attention and are therefore assumed to exhibit HSBs (H= 1).(5,7) Although this scenario seems unlikely in real-world settings, where these behaviors may only modify the probability of seeking medical care, classical TND studies could still offer better control for confounding by HSBs than other observational study designs such as cohorts or traditional case-control studies.(7,8)

To illustrate this point, **Figure 1(a)** shows a DAG that represents an ideal scenario for a classical TND study evaluating VE against medically attended and laboratory-confirmed symptomatic COVID-19. By “ideal”, we mean that the study is conducted in a population where HSBs are deterministic and only perfect tests are used (with 100% sensitivity and specificity). In this study, the effect of interest is represented by the path 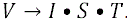. The square shapes of the nodes *S*, *T*, and *H* indicate that the study sample is restricted to symptomatic and tested individuals who sought medical care and therefore exhibit HSBs (*S* = 1, *T* = 1, *H* = 1). Conditioning on HSBs is critical because it not only allows control of confounding through the path 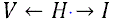 but also blocks other biasing paths (e.g., 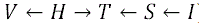) that were opened by conditioning on colliders such as testing (*T*). In other words, restricting the study sample in a classical TND study to symptomatic and tested individuals is essential to prevent (or minimize) confounding and collider-stratification bias by HSBs,(7) and thus, to identify the *RR_COVID_*.

**Figure 1:** Causal-directed acyclic graphs for test-negative-design (TND) studies of COVID-19 vaccine effectiveness: a) the classical TND setting, b) the alternative TND setting, c) potential for differential misclassification of outcome status in alternative TND studies. **Abbreviations**: *C*: Confounders; *H*: health and healthcare-seeking behaviors; *I*: Infection status; *Ix*: Measured infection status; *S*: COVID-19-like symptoms; *T*: SARS-CoV-2 diagnostic tests; *V*: COVID-19 vaccination status. **Notes**: The square nodes indicate that the variable is controlled by either the study design or the analysis, while unconditioned nodes are represented as circles.

### The case of the alternative TND

#### Potential for collider stratification bias and uncontrolled confounding by HSBs

Having studied the classical TND, we now turn to the challenges presented by the alternative approach. In this scenario, since the study sample includes all tested individuals, regardless of symptoms, the target parameter is no longer the *RR_COVID_*. Instead, it is the conditional RR for SARS-CoV-2 infection relative to vaccination status (*RR_infect_*), represented by the path 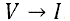. Mathematical definitions for *OR_infect_* and *RR_infect_* are given in **Table 3**.

To assess the identifiability of this target parameter, it is crucial to identify the specific drivers of testing leading to selection. Importantly, these drivers may have varied across settings, depending on the stage of the pandemic and the prevailing indications for testing.(39,40) For example, in a prospective, nonprobability-based, cross-sectional online survey of testing practices conducted between August 23, 2021, and March 12, 2022, among 418,279 U.S. adults aged ≥18 years, the most common reasons for testing, other than having symptoms, were exposure to COVID-19 (23.1%), a prerequisite for travel (20.6%), and requirement for work or school (16.1%).(39) Given that some of these reasons were mandatory at various points in time, we can no longer assume that the study sample is limited to healthcare seekers,(21) and that the TND effectively controls for bias related to HSBs. This may also be relevant to some hospital-based TND studies, as some institutions implemented policies for universal screening on admission.(41,42)

We illustrate this point in the DAG shown in **Figure 1(b)**, which represents an alternative TND study of VE against SARS-CoV-2 infection. In this DAG, the circular shapes of the nodes HSBs (H) and COVID-19-like symptoms (S) indicate that we are no longer conditioning on these variables. As a result, several biasing paths are opened, leading to uncontrolled confounding by HSBs (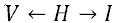; path 1 in **Figure S1**) and collider stratification bias (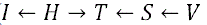 (path 2); 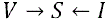 (path 3); 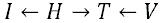 (path 4); 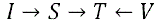 (path 5); and 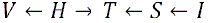 (path 6)). In other words, the *RR_infect_* is not identifiable in the alternative TND setting.

#### Potential for differential outcome misclassification

Another perceived benefit of the TND is its potential to reduce outcome misclassification compared to traditional cohort or case-control studies.(7,8) This is primarily attributed to the restriction of the study sample to individuals with confirmed infection status.(7,8) However, the inclusion of asymptomatic individuals in alternative TND studies could potentially compromise this apparent benefit and instead lead to differential misclassification of the outcome relative to the exposure status. The main reason for this is that while SARS-CoV-2 diagnostic tests have excellent specificity for diagnosing acute infection, their sensitivity is likely modified by the presence of symptoms.(40) For example, two studies comparing the diagnostic accuracy of SARS-CoV-2 tests, reported antigen test sensitivities in symptomatic individuals of 72.0% (95% confidence interval [95% CI], 63.7% to 79.0%) and 58.1% (95% CI, 40.2% to 74.1%) in those asymptomatic,(43) and nucleic acid amplification tests (NAATs) sensitivities in symptomatic individuals of 97.1% (95% CI, 96.7% to 97.3%) and 87.6% (95% CI, 85.2% to 89.6%) in those without symptoms.(44)

This concept is illustrated in the DAG shown in **Figure 1(c)**. This DAG introduces a new node for the measured infection status (*1x*), which may differ from its true value (*1*). There is differential misclassification of the outcome (*1*) relative to the exposure status (*V*) because, as described in **Table 2**, COVID-19 vaccines (*V*) may affect the presence of symptoms (*S*),(31–33) which in turn modify the sensitivity of SARS-CoV-2 tests (represented by the path 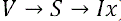). This situation may be further complicated when considering that the selection of a specific test and its respective diagnostic performance, may depend on a number of additional factors.(40) These include the intended use of the test (i.e., screening versus confirmation, with molecular tests preferred for confirming diagnoses of acute infection in symptomatic individuals(30,40)), the time of infection onset,(45) the stage of the pandemic(40), and the target population (e.g., travelers vs. frontline workers(40)). As a result, it is difficult to accurately predict the magnitude and direction of this bias without a comprehensive understanding of the causal structure underlying a given study.(46)

## Simulation study

### Methods

#### Overview

We conducted a simulation study to estimate and compare the magnitude of the bias in *OR_COVID_* and *OR_infect_* estimates — relative to their respective target parameters (*RR_COVID_* and *RR_infect_*) — under scenarios where a classical TND is considered valid. For simplicity, all simulations assume causal consistency,(38) no interference,(38) positivity,(38) and no unmeasured confounding other than that related to HSBs. To assess the bias pathways, we divided the simulation study into two parts. First, we evaluate the potential for collider bias and uncontrolled confounding by HSBs, assuming perfect diagnostic tests. Second, we incorporate the potential for differential outcome misclassification by simulating an extreme scenario where symptomatic individuals are tested exclusively with NAATs, and asymptomatic individuals are tested exclusively with antigen tests. Variables other than SARS-CoV-2 infection are assumed to be perfectly measured in all simulations.

#### Data generation process

To generate realistic data consistent with the DAG shown in **Table 2**, we generated 1,000 datasets (each with 1,000,000 observations) that simulated a cumulative prevalence of SARS-CoV-2 infection of ∼12.2%,(47) including an asymptomatic infection prevalence of ∼35%,(34) and a cumulative prevalence of fully vaccinated individuals of ∼54.6%.(48,49) In addition, we simulated the proportion of individuals who reported using any SARS-CoV-2 diagnostic test (either NAATs or antigen tests) within 30 days in the aforementioned survey (∼27.7%).(39) Each node in the DAG was generated conditionally on its parent nodes using the models shown in **Table 4**, with parameters for these models selected based on the available literature whenever possible (**Table S1**). Importantly, although the simulations were designed to represent an outpatient setting, similar results can be expected for hospital-based TND studies, as the data generating structure may be similar for both scenarios (**Table 2**).

**Table 4.**
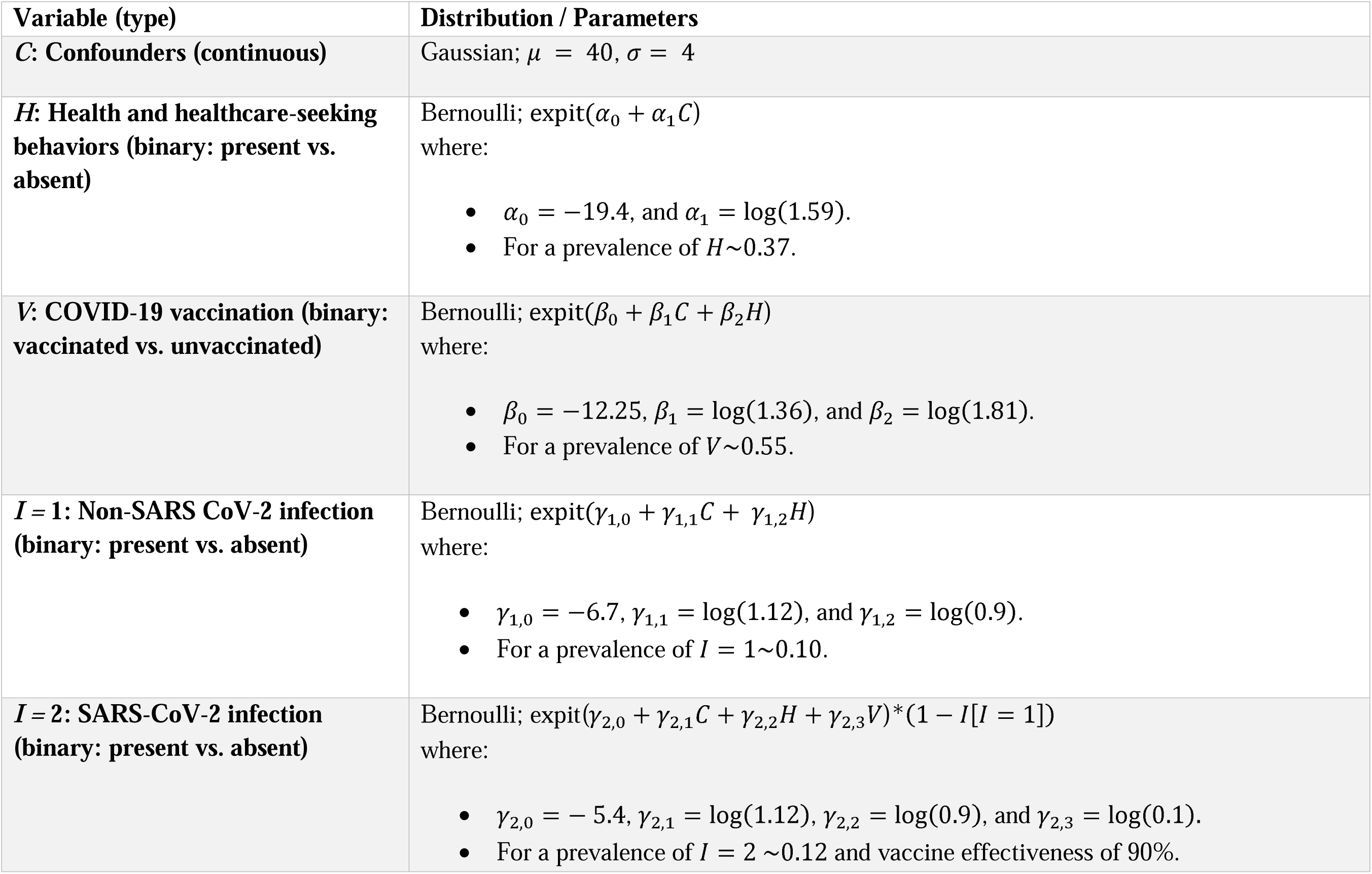
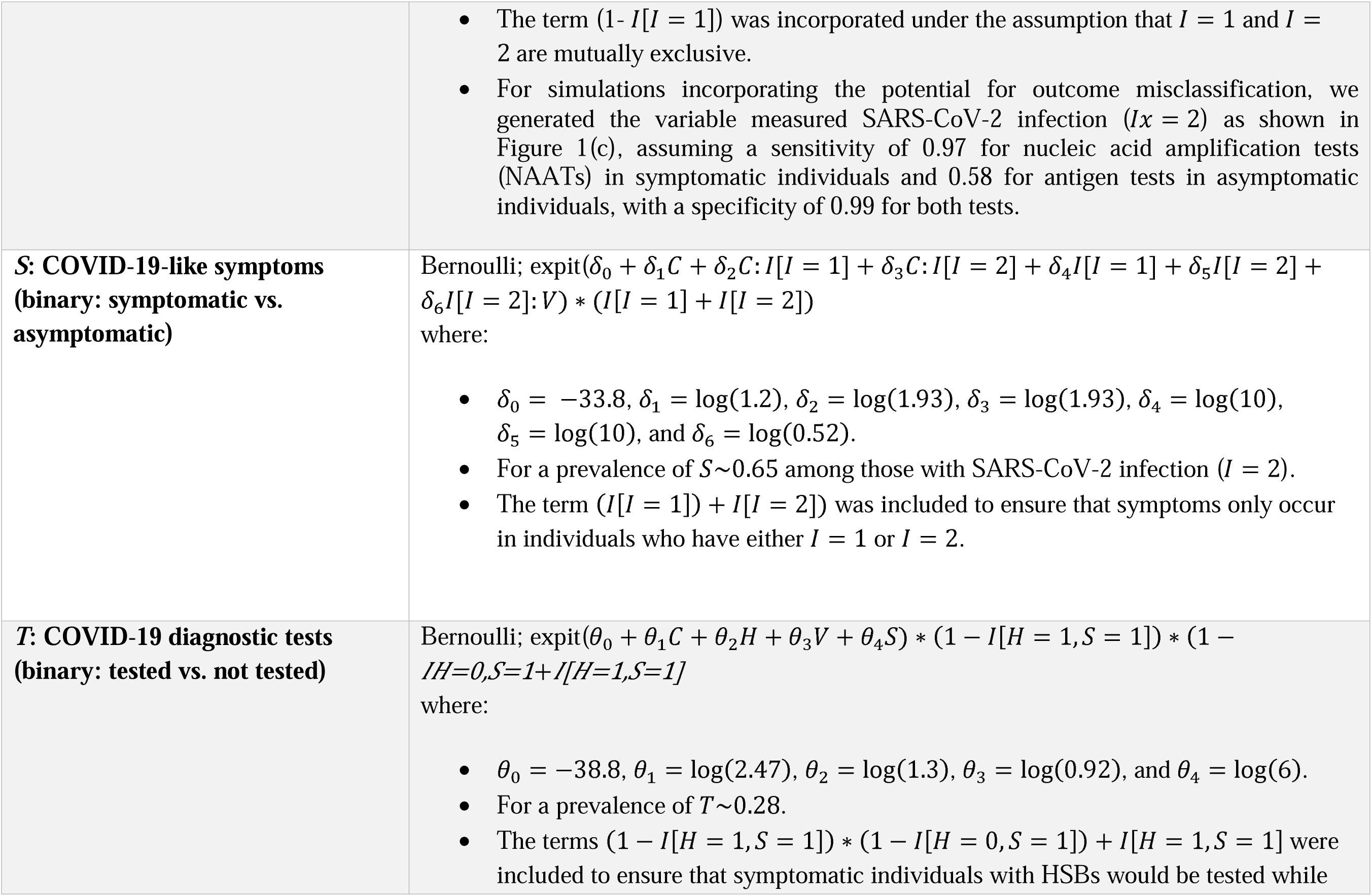
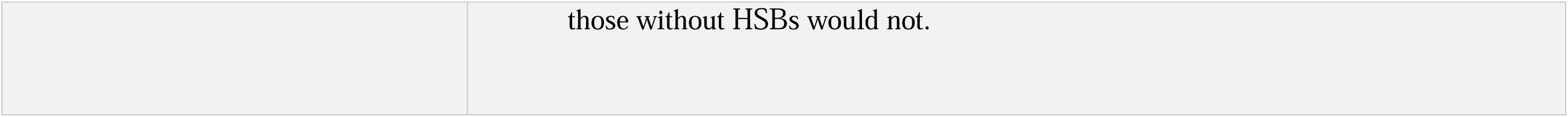
Models used in the simulation study to generate data consistent with the directed acyclic graph shown in Table 2.

#### Data analysis

After generating the data, we first created classical and alternative TND samples from each simulated dataset by selecting either symptomatic and tested individuals or all tested individuals, respectively. Second, we estimated the *OR_COVID_* (in the classical samples) and the *OR_infect_* (in the alternative samples) using logistic regression models conditional on the set of measured confounders (c). For these models, we used the true SARS-CoV-2 infection status as the outcome variable when evaluating the potential for collider bias and uncontrolled confounding by HSBs, and the measured SARS-CoV-2 infection status, when incorporating the potential for differential outcome misclassification. Third, to estimate the true value of the target parameters (*RR_COVID_* and *RR_infect_*), we generated two counterfactual populations, each consisting of 100,000,000 individuals, representing scenarios in which everyone was either vaccinated or unvaccinated. Finally, we estimated the bias by subtracting the true values of the target parameters from the exponentiated mean of the log(OR) estimates obtained from the 1,000 simulated datasets (denoted as 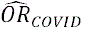 or 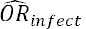. Additionally, we report the Monte Carlo standard error (MCSE, the standard deviation of the estimated log(ORs)) and the average standard error (aSE, the average of the standard error estimates of the log(ORs)).

To investigate the potential for larger biases, we iterated over the processes described above, each time varying the assumed strength of the relationships between the simulated variables. Specifically, we varied the magnitude of the association between *H* and *V* (parameter *β*_2_ in **Table 4**), between *H* and both *1*= 1 and *1*= 2 (parameters γ_l,2_ and γ_2,2_, respectively), as well as the relationships of *1*= 1 and *1*= 2 with *S* (δ_4_ and δ_S_, respectively). We also varied the coefficient for the interaction term of *1*= 2 and *V* with *S* (δ_6_), and the strengths of the relationships between *H* and *T* (*θ*_2_), between *V* and *T* (*θ*_3_), and between *S* and *T* (*θ*_4_). We adjusted these parameters according to the direction of the association between the independent and dependent variables. Specifically, for parameters representing a positive association (i.e., OR > 1), the strength was increased by units of 1 from 1.5 to 10.5. Conversely, for parameters representing a negative association (i.e., OR < 1), the strength was decreased from 0.95 to 0.05 by units of 0.1. We also varied the intercepts for *T* (*θ*_O_) and *1*= 2 (γ_2,O_), aiming for a baseline prevalence of testing from 0.25 to 0.7 and for a baseline prevalence of acute SARS-CoV-2 infection from 0.05 to 0.5. Finally, we simultaneously varied the strength of two, and then three, of the most influential parameters (i.e., *β*_2_, *β*_3_, and the intercept for *1*= 2). It should be noted that any adjustment to these parameters could lead to changes in the proposed prevalences for all simulated variables.

All analyses were performed using R (version 4.2.2, R Foundation for Statistical Computing, Vienna, Austria, 2022).(50)

### Results

When we assessed the potential for collider bias and uncontrolled confounding by HSBs we found a “true value” for *RR_COVID_* of 0.149 and an estimated 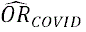 of 0.141 (MCSE and aSE = 0.019; bias = –0.008). On the other hand, the “true value” for *RR_infect_* was 0.142, with an estimated 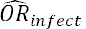 of 0.125 (MCSE and aSE = 0.014; bias = –0.016). The results after varying the strength of selected parameters for this scenario, either individually or simultaneously, are shown in **Figures 2**-**3** and **Tables S2**-**S3**. In every case, the classical TND outperformed the alternative in terms of bias, with the biases for 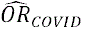 and 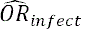 ranging from –0.034 to –0.001 and from –0.211 to 0.728, respectively. When we incorporated imperfect tests, we found estimates for 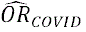 of 0.154 (MCSE = 0.019; aSE = 0.018; bias = 0.005) and 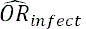 of 0.146 (MCSE and aSE = 0.014; bias = 0.004), respectively. However, after varying the strength of selected parameters for this scenario, either individually or simultaneously, we found larger bias for 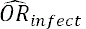 (ranging from –0.188 to 0.766) than for 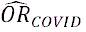 (ranging from –0.005 to 0.018) (**Figures 4-5** and **Tables S4-S5**).

**Figure 2:** Bias in odds ratio (OR) estimates for symptomatic COVID-19 (**OR_COVID_**) and SARS-CoV-2 infection (**OR_infect_**) relative to their respective target parameters (**RR_COVID_** and **RR_infect_**) after varying the strength of selected parameters (assuming perfect SARS-CoV-2 diagnostic tests). **Abbreviations**: *C*: Confounders; *H*: health and healthcare-seeking behaviors; *I*: Infection status; *I* =-2: SARS-CoV-2 infection status; OR, odds ratio; RR, risk ratio; *S*: COVID-19-like symptoms; *T*: SARS-CoV-2 diagnostic tests; TND, test-negative design; V: COVID-19 vaccination status. **Notes**: Bias of the OR = (exp(mean(measured log[OR])) – (true RR).

**Figure 3:** Bias in odds ratio (OR) estimates for symptomatic COVID-19 (**OR_COVID_**) and SARS-CoV-2 infection (**OR_infect_**) relative to their respective target parameters (**RR_COVID_** and **RR_infect_**) after simultaneously varying the strength of two selected parameters (assuming perfect SARS-CoV-2 diagnostic tests). **Abbreviations**: *H*: health and healthcare-seeking behaviors; *I* =-2: SARS-CoV-2 infection status; NAATs, Nucleic Acid Amplification Tests; OR, odds ratio; RR, risk ratio; *T*: SARS-CoV-2 diagnostic tests; TND, test-negative design; V: COVID-19 vaccination status. **Notes**: Bias of the OR = (exp(mean(measured log[OR])) – (true RR).

**Figure 4:** Bias in odds ratio (OR) estimates for symptomatic COVID-19 (**OR_COVID_**) and SARS-CoV-2 infection (**OR_infect_**) relative to their respective target parameters (**RR_COVID_** and **RR_infect_**) after varying the strength of selected parameters (assuming symptomatic individuals are tested exclusively with NAATs and asymptomatic individuals are tested exclusively with antigen tests). **Abbreviations**: *C*: Confounders; *H*: health and healthcare-seeking behaviors; *I*: Infection status; *I* =-2: SARS-CoV-2 infection status; NAATs, Nucleic Acid Amplification Tests; OR, odds ratio; RR, risk ratio; *S*: COVID-19-like symptoms; *T*: SARS-CoV-2 diagnostic tests; TND, test-negative design; V: COVID-19 vaccination status. **Notes**: Bias of the OR = (exp(mean(measured log[OR])) – (true RR).

**Figure 5:** Bias in odds ratio (OR) estimates for symptomatic COVID-19 (**OR_COVID_**) and SARS-CoV-2 infection (**OR_infect_**) relative to their respective target parameters (**RR_COVID_** and **RR_infect_**) after simultaneously varying the strength of two selected parameters (assuming symptomatic individuals are tested exclusively with NAATs and asymptomatic individuals are tested exclusively with antigen tests). **Abbreviations**: *H*: health and healthcare-seeking behaviors; *I* =-2: SARS-CoV-2 infection status; NAATs, Nucleic Acid Amplification Tests; OR, odds ratio; RR, risk ratio; T: SARS-CoV-2 diagnostic tests; TND, test-negative design; *V*: COVID-19 vaccination status. **Notes**: Bias of the OR = (exp(mean(measured log[OR])) – (true RR).

## Discussion

The TND has become a preferred method for assessing VE against pathogens such as influenza, largely due to its seamless integration with existing laboratory-based surveillance systems.(6) More recently, this observational study design has been widely used to estimate VE against COVID-19.(21) This widespread application was made possible primarily by the extensive use of SARS-CoV-2 testing during the pandemic, which generated large amounts of potential data for TND studies.(21) However, the use of these data for VE estimation poses unique validity challenges.(21)

Originally, the TND was proposed as an efficient way to identify a control group (test-negative controls) for individuals who contracted the vaccine-targeted infection (test-positive cases) while also accounting for confounding by HSBs.(5,7,8) However, to effectively control for such bias in TND studies, researchers must assume that individuals seeking care and undergoing testing have comparable HSBs.(5,7,8) In the pre-COVID-19 era, testing for diseases such as influenza was primarily driven by clinical indications and focused on those who actively sought care for symptoms.(51) This arguably made it easier for researchers to assume that individuals enrolled in TND studies had comparable HSBs, provided the same clinical definition was used for cases and controls. However, this paradigm shifted with COVID-19, when testing criteria were expanded to identify asymptomatic infections, often on a “mandatory” basis, in order to break transmission chains.(40) In this article, we have shown that TND applications that ignore this fact and include individuals regardless of their symptoms (i.e., using the alternative TND) can potentially introduce collider stratification bias, uncontrolled confounding by HSBs, and differential misclassification of the outcome relative to the exposure status.

Our simulation study, based on educated guesses for parameter values and representing a scenario in which the classical TND would yield unbiased estimates for its target parameter, showed only minor bias in the alternative TND setting. However, we also showed that the inclusion of asymptomatic individuals can lead to substantial bias in some scenarios. For example, our simulations showed that as the effect of HSBs on vaccination increases (OR > 1), the estimated 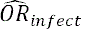 can become progressively closer to the null relative to its target parameter, *RR_infect_*, leading to further underestimation of VE when expressed as 1 – OR × 100. Similarly, in settings where vaccination status is more negatively correlated with testing (OR < 1), the 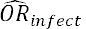 would also be progressively biased towards the null. In addition, we observed that a higher baseline prevalence of SARS-CoV-2 infection would lead to a greater bias away from the null of 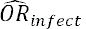, resulting in an overestimation of VE. We also found that these biases may be exacerbated by the introduction of differential outcome misclassification when different diagnostic approaches are used for symptomatic and asymptomatic individuals. Of note, the TND is well known to be particularly susceptible to misclassification bias compared to cohort and traditional case-control studies.(52) It is also important to mention that if one were to condition on the reason for testing or restrict the study sample to asymptomatic individuals, we would expect to observe the same bias described here, as all biasing pathways would remain open.

It could be argued that the biases discussed here may be only relevant to retrospective TND studies that rely on routinely collected health data (i.e., data collected without predetermined research questions(53)). For example, some TND studies using administrative data sources may have included asymptomatic individuals simply because of a lack of information on reasons for testing, as acknowledged by some authors.(14,15) However, it should be noted that some retrospective TND studies of COVID-19 VE intentionally included asymptomatic individuals to estimate VE against infection, even when data on symptoms were available.(10–12,16,17) In fact, some prospective TND studies (i.e., those collecting primary research data) conducted during the COVID-19 pandemic also included individuals regardless of symptoms.(18–20) That said, the potential for inclusion of asymptomatic individuals may be influenced not only by the data source, but also by the testing practices in the specific context or time frame in which the study is conducted.

The changing landscape of COVID-19 testing practices throughout the pandemic further highlights the need for proper attention to context in TND studies. At the onset of the COVID-19 pandemic, testing was primarily NAAT-based and focused on symptomatic individuals.(40) However, as the pandemic progressed, testing guidelines were modified to support mass testing of asymptomatic individuals as a tool for pandemic containment.(40) Any TND study conducted under these circumstances, whether prospective or retrospective, would be at risk of the bias described here if a clinical definition was not included in the study’s eligibility criteria. Thus, we believe that the issues discussed here are relevant not only to TND studies using routinely collected health and administrative data, but also to all studies conducted in settings where the rationale for testing extends beyond purely clinical reasons. **Table 5** provides a list of suggested strategies to minimize bias in TND studies conducted in such scenarios.

**Table 5.**
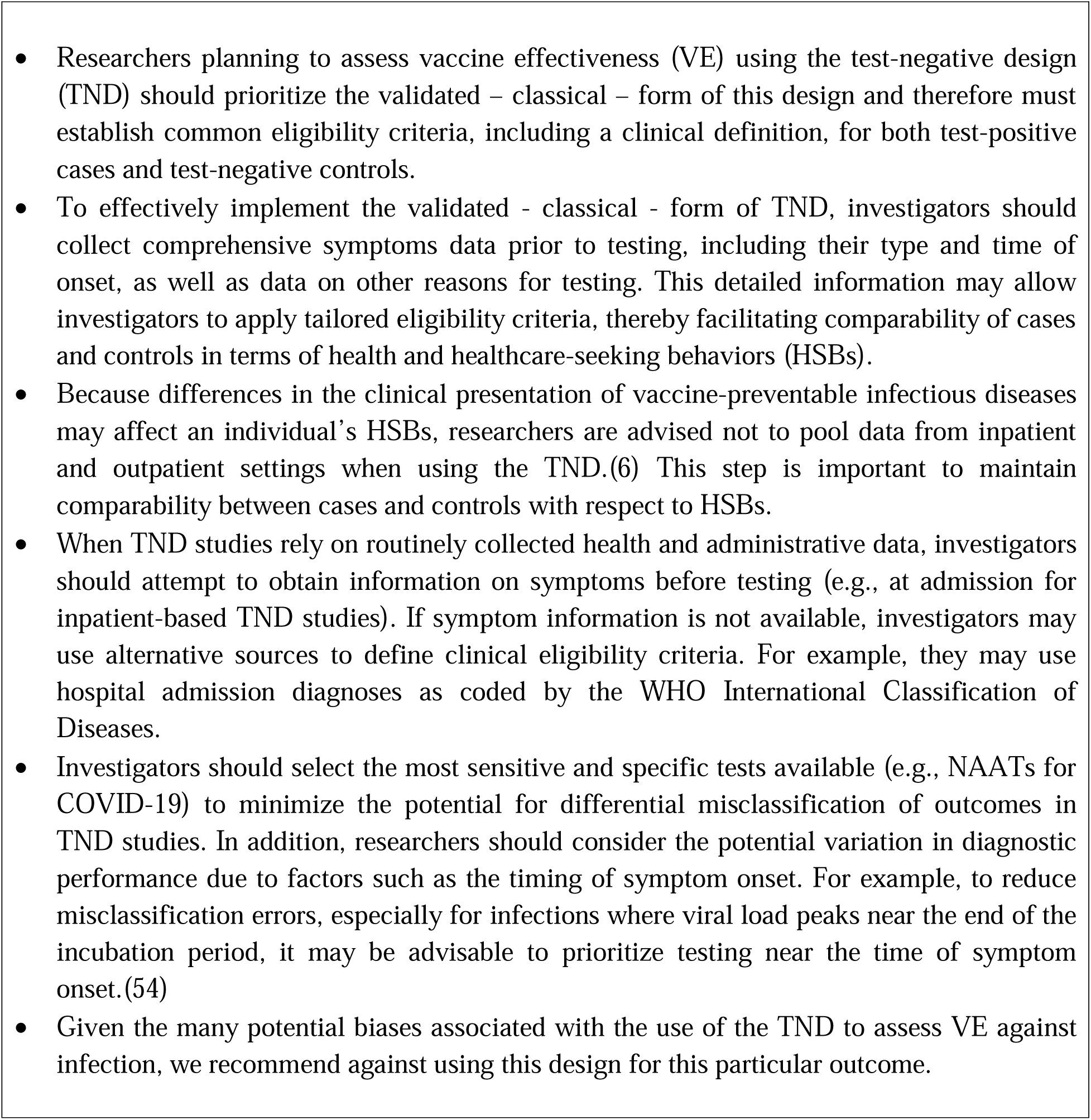
Suggested strategies for minimizing bias in TND studies conducted during infectious disease outbreaks.

It is worth noting that there may be some specific circumstances in which the magnitude of the biases discussed here could be attenuated by other features of the study design, particularly those that allow for the selection of a population with comparable HSBs. For example, TND studies focusing on booster effectiveness may be less susceptible to HSBs-related bias because subjects in these studies typically have completed a primary immunization schedule and are therefore expected to have more similar HSBs.(21) Likewise, some might argue that hospital-based TND studies are likely to include individuals with more homogeneous HSBs because only those evaluated in clinical settings are eligible. However, as discussed elsewhere,(7) it may be unrealistic to assume that all hospitalized individuals have the same levels of HSBs. In addition, it should be noted that not all SARS-CoV-2 tests performed in hospitals under pandemic conditions were triggered by COVID-19-like symptoms; for example, some institutions mandated universal screening on admission.(41,42) Consequently, hospital-based TND studies that include all individuals tested regardless of the reason for testing,(13,55) would be expected to have biases similar to those discussed here.

This study has numerous strengths but also carries limitations. Among the strengths, our simulation study was illustrated by DAGs, which helps to clarify the potential sources of bias. In addition, while this article focuses on the COVID-19 setting, the issues discussed here are also relevant to TND studies using the alternative approach in the context of other infectious diseases. This may be particularly true in scenarios of widespread testing, such as during epidemics or pandemics, where testing protocols and preferences are influenced by clinical and public health considerations, or in situations where self-testing for various infections is available.

In terms of limitations, the strength of the relationships depicted in the DAG may vary. For example, the path between vaccination and testing (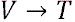) may be weak in some contexts. Nevertheless, collider bias could still occur through these nodes because unobservable factors, including HSBs, could confound the association between *V* and *T*. Another limitation lies in the values assigned to the parameters in our simulations. Although we relied on existing literature, these values are still estimates. However, varying these parameter values supported our theory-based hypothesis that relevant biases may occur in some settings. Finally, our study focused on specific issues related to the inclusion of asymptomatic individuals in TND studies. However, we did not assess additional complexities, such as the possibility of differential measurement error in exposure, outcome, or covariate status based on testing rationale, or the nuances in data quality and challenges specific to their respective sources, such as clinical versus other settings.(21)

## Conclusions

In conclusion, the inclusion of asymptomatic individuals in TND studies of COVID-19 VE may lead to collider stratification bias, uncontrolled confounding by HSBs, and differential misclassification of the outcome relative to exposure status. Researchers designing or applying the TND for VE estimation need to be aware of these potential biases. Further research is needed to identify additional design or analytic strategies to control for biases related to HSBs that may improve the validity and utility of the TND for estimating VE against infection.

## Acknowledgments

A part of this work was presented at the Society for Epidemiologic Research (SER) conference held in Portland in June 2023, specifically during the oral abstract session entitled “Study Design – It’s All About the People”. A preprint version of this manuscript was published on medRxiv on November 16, 2023, and is available at https://doi.org/10.1101/2023.11.16.23298633.

## Disclaimer

The views expressed in this article are those of the authors and do not necessarily reflect the official position of any agency of the authors’ institutions or funders.

## Data Availability Statement

The code and a sample dataset can be found at the following link: https://github.com/ortizbrizuela/

## Supplementary Material

## Supplementary Text

## Appendix S1: A brief overview of causal directed acyclic graph theory and terminology

A causal Directed Acyclic Graph (DAG) is a collection of “nodes” representing random variables, with their causal (temporal) relationships illustrated by unidirectional (directed) arrows, also known as “arcs.”^1-3^ In a DAG, no variable can be caused by itself, either directly or indirectly through other variables, making it “acyclic.” When an arc connects two nodes, the variable from which it originates is called the “parent,” and the variable to which it points is referred to as its “child.” Moreover, a “path” is any sequence of arcs connecting two nodes, regardless of their direction. A “directed path” is a path where all arcs point in the same direction, representing a causal path. In contrast, non-directed paths between two variables that lead to their association (as explained below) are called “biasing paths”. For example, the path ***A ← B → C*** is a non-directed (and biasing) path between *A* and *C*, also known as a “backdoor path” because it starts with an arc pointing to A and ends with an arrow pointing to *C*.

In a directed path, variables in proximal positions are called “ancestors” of those in distal positions, and those in distal positions are “descendants” of those in proximal positions. Furthermore, nodes in any path can be classified as “colliders” or “non-colliders.” A “collider” in a path is a node with its preceding and subsequent arcs pointing at it (e.g., ***B*** is a collider in the path ***A → B ← C***). In contrast, a “non-collider” can be classified as a “mediator” if the variable occupies an intermediate position in the directed path, or as a “fork” if the preceding and subsequent arcs emerge from it. For example, ***B*** is a mediator in the path ***A → B → C***, while ***B*** is a fork in the path ***A ← B → C***.

One of the main benefits of DAGs is that they allow users to evaluate potential sources of association between variables and, consequently, to identify possible sources of bias. In general, according to DAG theory, two nodes are expected to be statistically associated if any path between them is “open” (in this case, variables are categorized as “d-connected,” otherwise they are considered “d-separated”).^1,3^ A path between two nodes is “closed” if 1) it contains a collider (for which the analysis has not been conditioned on, nor on its descendants), or 2) if we condition our analysis on any non-collider within the path.^1^ For example, “confounding” (i.e., the presence of open backdoor paths) or “collider bias” (i.e., opening a non-causal path by conditioning on a collider or its descendants) are examples of sources of non-causal (biased) associations between two variables that can be detected using DAGs.

Importantly, in order to attribute any found association between two variables in the DAG solely to open paths, we must assume that the DAG is missing no variable that affects two or more variables in it, that no variable that affects selection or is used for stratification that may be a collider is missing, and that there is no measurement or random error that could explain the association.^3^ We refer readers to other sources for a more comprehensive yet gentle introduction to DAG theory.^1-3^

## Appendix S2: Identifiability of causal effects under outcome-dependent sample selection

In studies where sampling depends on outcome status, such as traditional case-control studies or the test-negative design (TND), the risk of the outcome, the risk ratio, and the risk difference are subject to bias.^4,5^ Didelez et al.^6^ outlined two conditions that can be evaluated graphically using directed acyclic graphs (DAGs) necessary for both identifying and generalizing the conditional causal odds ratio (OR) in such circumstances:

1. The first condition — needed to identify the conditional causal OR within the study sample — requires that no backdoor path remains open between exposure and outcome (see **Appendix S1** for further details on DAG theory). This means that in the classical TND, after conditioning on health and healthcare-seeking behaviors (HSBs (H)) and the set of measured confounders (C), no backdoor path must remain open between the node vaccination status (V) and all components of the outcome medically attended and laboratory-confirmed symptomatic COVID-19 (/· S · T). This condition can be achieved within the classical TND by restricting the study sample to healthcare seekers (H = 1) and conditioning on the set of measured confounders (C) during the analysis. On the other hand, for this condition to be met in the alternative TND, no backdoor path must remain open between the vaccination status (V) and infection (/) after conditioning on HSBs (H) and the set of measured confounders (C). However, because the study sample is not limited to individuals with HSBs, it may not be possible to achieve this condition in the alternative TND. Therefore, while the conditional causal OR for medically attended and laboratory-confirmed symptomatic infection can be identified in the classical TND, the conditional causal OR for infection may remain unidentified in the alternative TND (our manuscript discusses numerous instances where conditional exchangeability on exposure is violated in the alternative TND).
2. The second condition — needed to generalize the conditional causal OR from the study sample to the entire population — requires the node for exposure status to be independent of the node selection, after conditioning on the outcome and a set of measured variables. In the classical TND, this means that the node representing an individual’s vaccination status () must be independent of the node representing selection into the study (, see DAG below), after conditioning on the outcome medically attended and laboratory-confirmed symptomatic COVID-19 (), HSBs (), and the set of measured confounders (). Using notation:. In the alternative TND, this condition translates into the node representing an individual’s vaccination status () being independent of the node selection into the study (), after conditioning on infection (), HSBs (), and the set of measured confounders (). Using notation: . In other words, based on our assumed DAG, only the conditional causal OR for medically attended and laboratory-confirmed symptomatic infection can be identified and generalized in the classical TND. Conversely, in the alternative TND, the conditional causal OR for infection may not be identifiable or generalizable to the broader population.

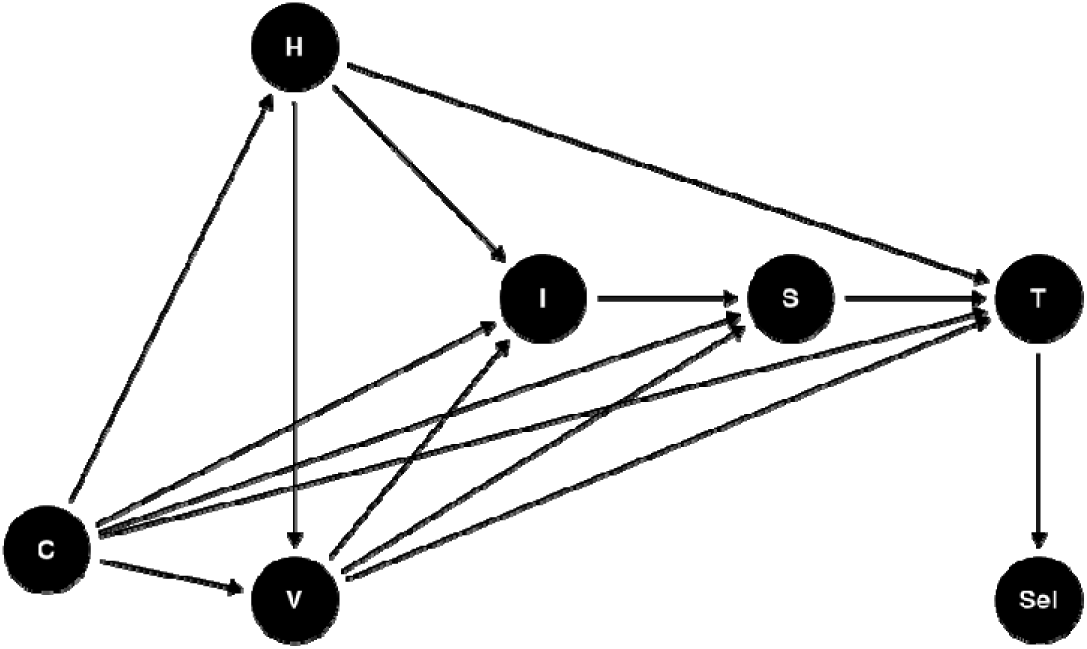

**Abbreviations**: *C*: Confounders; *H*: health and healthcare-seeking behaviors; *I*: Infection status; *Ix*: Measured infection status; *S*: COVID-19-like symptoms; *Sel*: selection into the study sample; *V*: COVID-19 vaccination status; *T*: SARS-CoV-2 diagnostic tests.

## Supplementary Tables

**Table S1:**
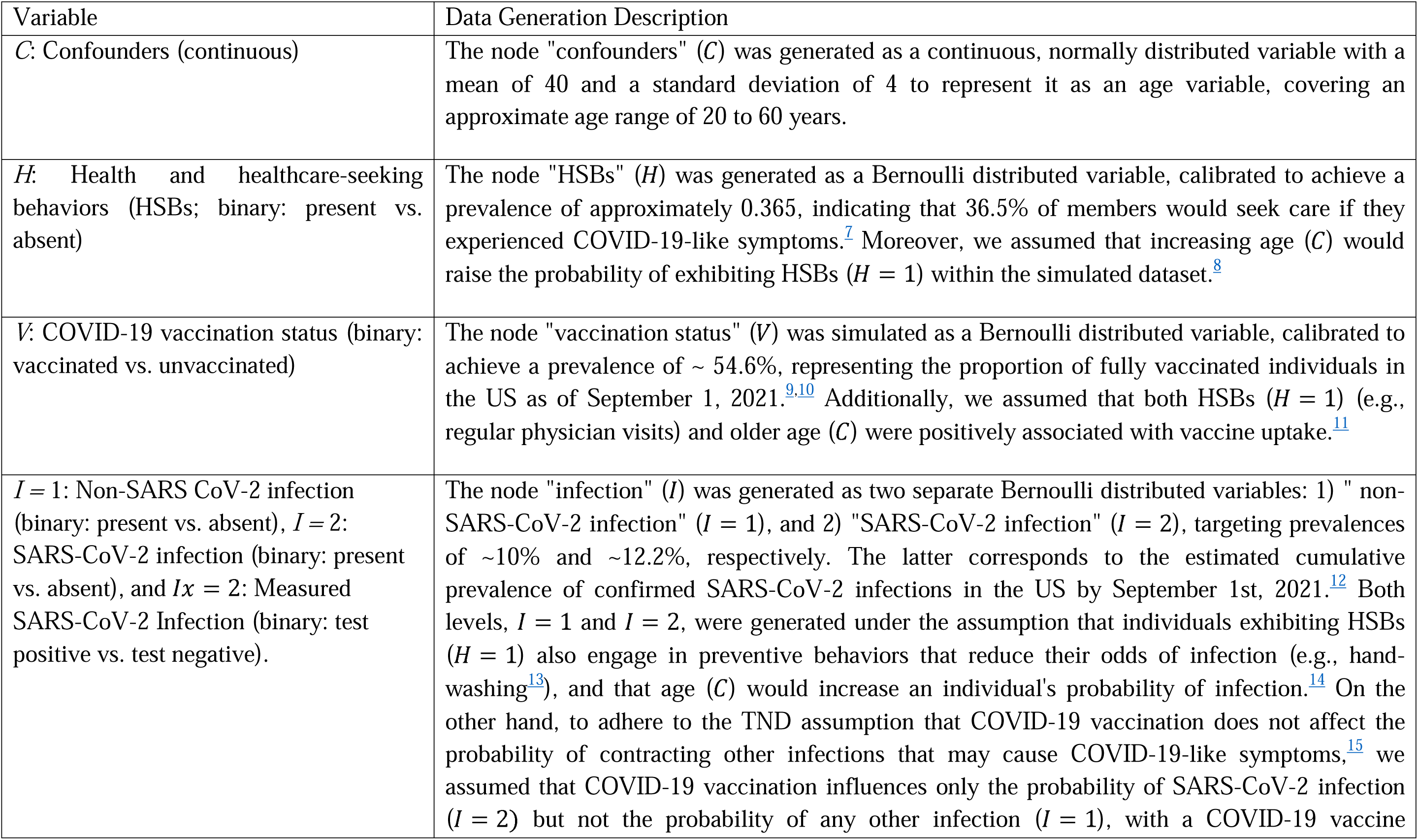
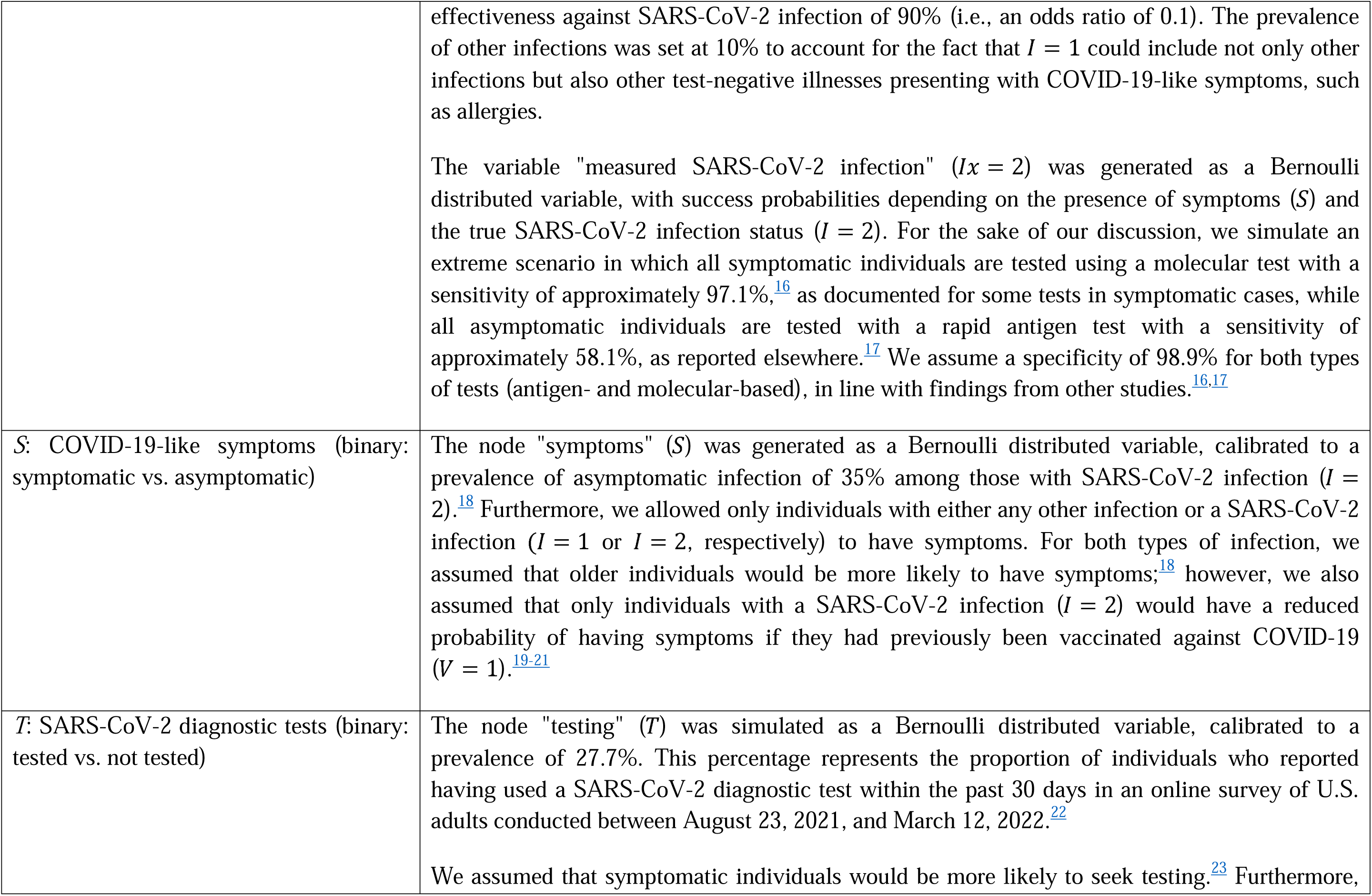
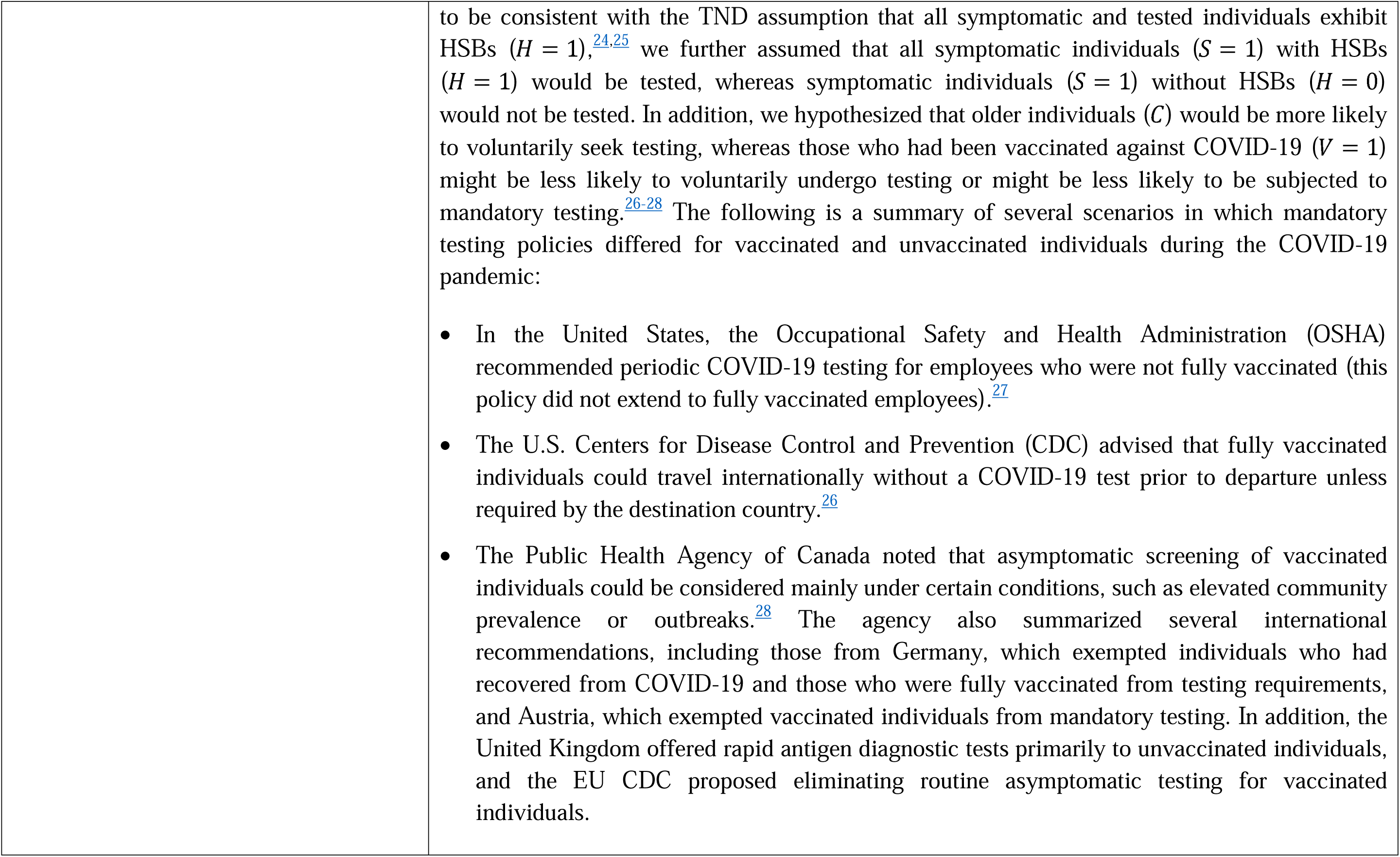
Data generation process for the DAG-guided simulation study.

**Table S2:**
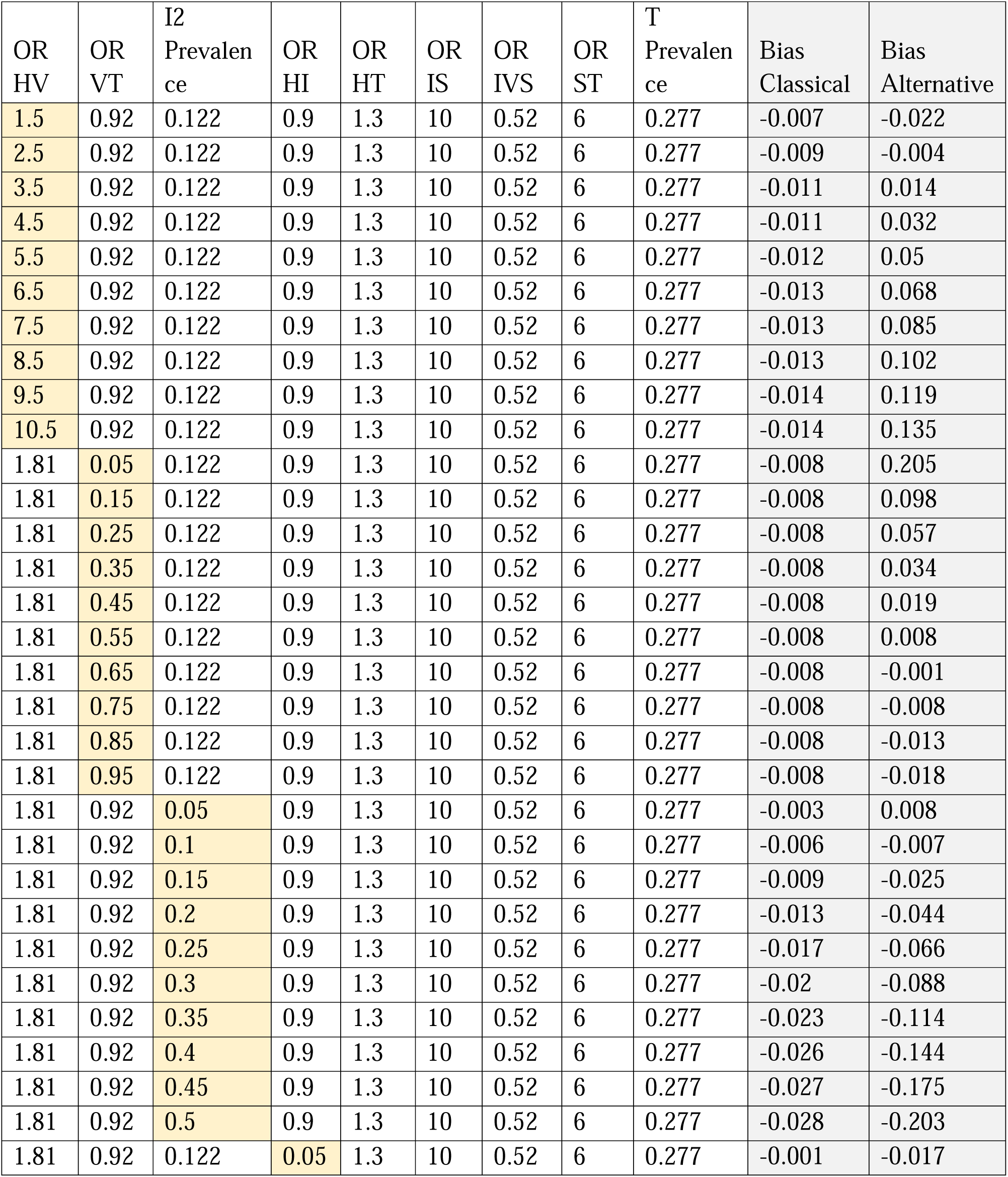
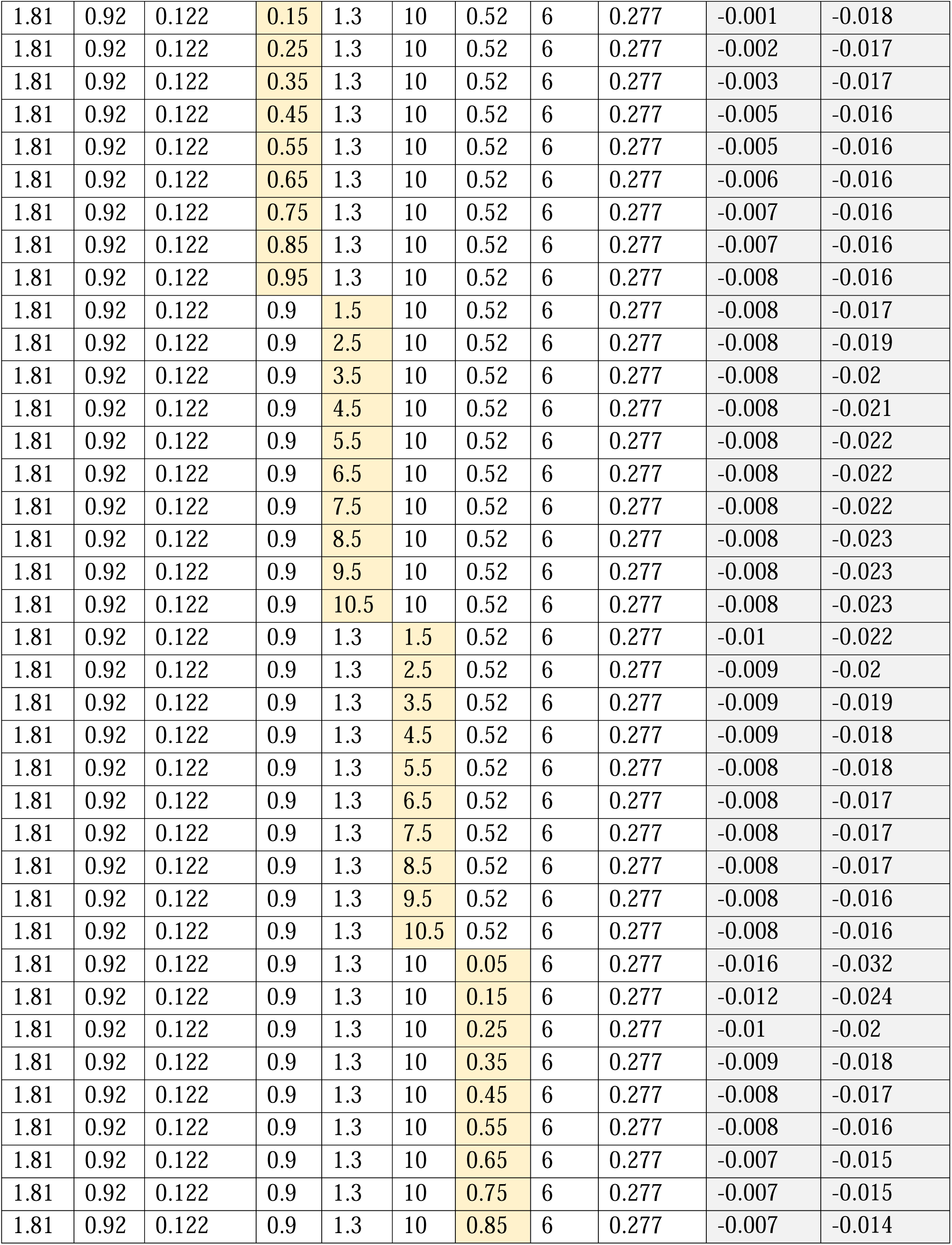
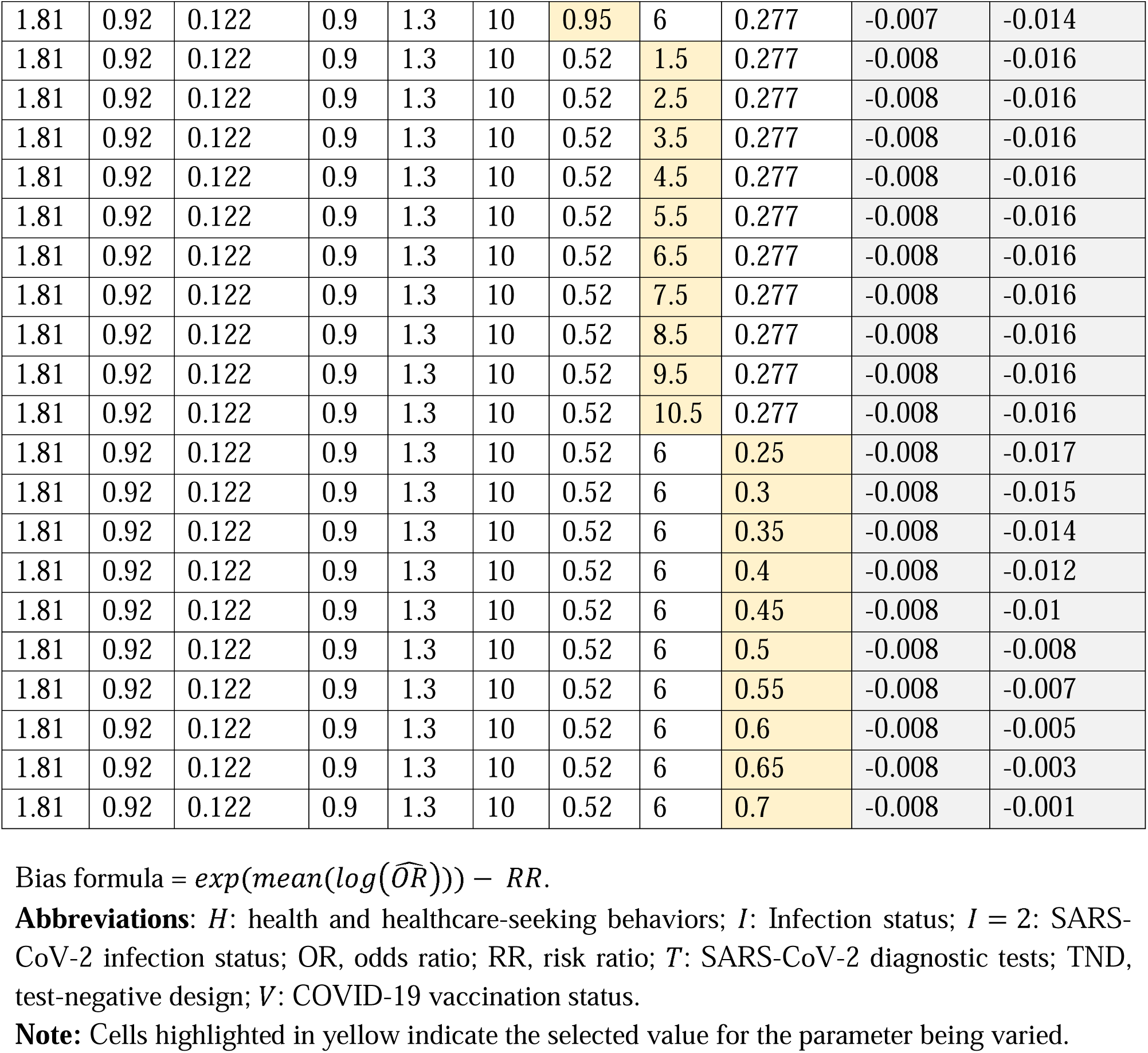
Bias in odds ratio (OR) estimates for symptomatic COVID-19 (*OR_COVID_*) and SARS-CoV-2 infection (*OR_infect_*) relative to their target parameters (*RR_COVID_* and *RR_infect_*, respectively) after individually varying the strength of one selected parameters (assuming perfect SARS-CoV-2 diagnostic tests).

**Table S3:** Bias in odds ratio (OR) estimates for symptomatic COVID-19 (*OR_COVID_*) and SARS-CoV-2 infection (*OR_infect_*) relative to their target parameters (*RR_COVID_* and *RR_infect_*, respectively) after simultaneously varying the strength of two or three selected parameters (assuming perfect SARS-CoV-2 diagnostic tests).

| OR HV | OR VT | I2 Prevalence | Bias Classical | Bias Alternative |
| --- | --- | --- | --- | --- |
| 1.5 | 0.95 | 0.122 | -0.007 | -0.023 |
| 2.5 | 0.85 | 0.122 | -0.009 | 0.000 |
| 3.5 | 0.75 | 0.122 | -0.011 | 0.026 |
| 4.5 | 0.65 | 0.122 | -0.011 | 0.056 |
| 5.5 | 0.55 | 0.122 | -0.012 | 0.091 |
| 6.5 | 0.45 | 0.122 | -0.013 | 0.135 |
| 7.5 | 0.35 | 0.122 | -0.013 | 0.192 |
| 8.5 | 0.25 | 0.122 | -0.013 | 0.272 |
| 9.5 | 0.15 | 0.122 | -0.014 | 0.403 |
| 10.5 | 0.05 | 0.122 | -0.014 | 0.699 |
| 1.5 | 0.92 | 0.5 | -0.025 | -0.209 |
| 2.5 | 0.92 | 0.45 | -0.032 | -0.163 |
| 3.5 | 0.92 | 0.4 | -0.034 | -0.114 |
| 4.5 | 0.92 | 0.35 | -0.032 | -0.067 |
| 5.5 | 0.92 | 0.3 | -0.029 | -0.024 |
| 6.5 | 0.92 | 0.25 | -0.025 | 0.016 |
| 7.5 | 0.92 | 0.2 | -0.020 | 0.055 |
| 8.5 | 0.92 | 0.15 | -0.016 | 0.092 |
| 9.5 | 0.92 | 0.1 | -0.011 | 0.129 |
| 10.5 | 0.92 | 0.05 | -0.006 | 0.164 |
| 1.81 | 0.95 | 0.5 | -0.028 | -0.205 |
| 1.81 | 0.85 | 0.45 | -0.027 | -0.170 |
| 1.81 | 0.75 | 0.4 | -0.026 | -0.131 |
| 1.81 | 0.65 | 0.35 | -0.023 | -0.094 |
| 1.81 | 0.55 | 0.3 | -0.020 | -0.060 |
| 1.81 | 0.45 | 0.25 | -0.017 | -0.026 |
| 1.81 | 0.35 | 0.2 | -0.013 | 0.010 |
| 1.81 | 0.25 | 0.15 | -0.009 | 0.050 |
| 1.81 | 0.15 | 0.1 | -0.006 | 0.105 |
| 1.81 | 0.05 | 0.05 | -0.003 | 0.218 |
| 1.5 | 0.95 | 0.5 | -0.025 | -0.211 |
| 2.5 | 0.85 | 0.45 | -0.032 | -0.157 |
| 3.5 | 0.75 | 0.4 | -0.034 | -0.099 |
| 4.5 | 0.65 | 0.35 | -0.032 | -0.039 |
| 5.5 | 0.55 | 0.3 | -0.029 | 0.022 |
| 6.5 | 0.45 | 0.25 | -0.025 | 0.087 |
| 7.5 | 0.35 | 0.2 | -0.020 | 0.164 |
| 8.5 | 0.25 | 0.15 | -0.016 | 0.263 |
| 9.5 | 0.15 | 0.1 | -0.011 | 0.412 |
| 10.5 | 0.05 | 0.05 | -0.006 | 0.728 |

Bias formula = $\exp(\text{mean}(\log(\widehat{OR}))) - RR$ .
**Abbreviations:** *H*: health and healthcare-seeking behaviors; *I*: Infection status; *I* = 2: SARS-CoV-2 infection status; OR, odds ratio; RR, risk ratio; *T*: SARS-CoV-2 diagnostic tests; TND, test-negative design; *V*: COVID-19 vaccination status.
**Note:** Cells highlighted in yellow indicate the selected value for the parameter being varied.

**Table S4:** Bias in odds ratio (OR) estimates for **measured** symptomatic COVID-19 (*OR_COVID_*) and **measured** SARS-CoV-2 infection (*OR_infect_*) relative to their target parameters (*RR_COVID_* and *RR_infect_*, respectively) after individually varying the strength of selected parameters (assuming symptomatic individuals are tested exclusively with NAATs and asymptomatic individuals are tested exclusively with antigen tests).

| OR<br>HV | OR<br>VT | I2<br>Prevalen<br>ce | OR<br>HI | OR<br>HT | OR<br>IS | OR<br>IVS | OR<br>ST | T<br>Prevalen<br>ce | Bias<br>Classical | Bias<br>Alternative |
| --- | --- | --- | --- | --- | --- | --- | --- | --- | --- | --- |
| 1.5 | 0.92 | 0.122 | 0.9 | 1.3 | 10 | 0.52 | 6 | 0.277 | 0.006 | -0.002 |
| 2.5 | 0.92 | 0.122 | 0.9 | 1.3 | 10 | 0.52 | 6 | 0.277 | 0.003 | 0.018 |
| 3.5 | 0.92 | 0.122 | 0.9 | 1.3 | 10 | 0.52 | 6 | 0.277 | 0.002 | 0.039 |
| 4.5 | 0.92 | 0.122 | 0.9 | 1.3 | 10 | 0.52 | 6 | 0.277 | 0.001 | 0.059 |
| 5.5 | 0.92 | 0.122 | 0.9 | 1.3 | 10 | 0.52 | 6 | 0.277 | 0 | 0.079 |
| 6.5 | 0.92 | 0.122 | 0.9 | 1.3 | 10 | 0.52 | 6 | 0.277 | 0 | 0.098 |
| 7.5 | 0.92 | 0.122 | 0.9 | 1.3 | 10 | 0.52 | 6 | 0.277 | 0 | 0.118 |
| 8.5 | 0.92 | 0.122 | 0.9 | 1.3 | 10 | 0.52 | 6 | 0.277 | -0.001 | 0.137 |
| 9.5 | 0.92 | 0.122 | 0.9 | 1.3 | 10 | 0.52 | 6 | 0.277 | -0.001 | 0.156 |
| 10.5 | 0.92 | 0.122 | 0.9 | 1.3 | 10 | 0.52 | 6 | 0.277 | -0.001 | 0.175 |
| 1.81 | 0.05 | 0.122 | 0.9 | 1.3 | 10 | 0.52 | 6 | 0.277 | 0.005 | 0.226 |
| 1.81 | 0.15 | 0.122 | 0.9 | 1.3 | 10 | 0.52 | 6 | 0.277 | 0.005 | 0.12 |
| 1.81 | 0.25 | 0.122 | 0.9 | 1.3 | 10 | 0.52 | 6 | 0.277 | 0.005 | 0.079 |
| 1.81 | 0.35 | 0.122 | 0.9 | 1.3 | 10 | 0.52 | 6 | 0.277 | 0.005 | 0.055 |
| 1.81 | 0.45 | 0.122 | 0.9 | 1.3 | 10 | 0.52 | 6 | 0.277 | 0.005 | 0.04 |
| 1.81 | 0.55 | 0.122 | 0.9 | 1.3 | 10 | 0.52 | 6 | 0.277 | 0.005 | 0.028 |
| 1.81 | 0.65 | 0.122 | 0.9 | 1.3 | 10 | 0.52 | 6 | 0.277 | 0.005 | 0.02 |
| 1.81 | 0.75 | 0.122 | 0.9 | 1.3 | 10 | 0.52 | 6 | 0.277 | 0.005 | 0.013 |
| 1.81 | 0.85 | 0.122 | 0.9 | 1.3 | 10 | 0.52 | 6 | 0.277 | 0.005 | 0.007 |
| 1.81 | 0.95 | 0.122 | 0.9 | 1.3 | 10 | 0.52 | 6 | 0.277 | 0.005 | 0.003 |
| 1.81 | 0.92 | 0.05 | 0.9 | 1.3 | 10 | 0.52 | 6 | 0.277 | 0.01 | 0.06 |
| 1.81 | 0.92 | 0.1 | 0.9 | 1.3 | 10 | 0.52 | 6 | 0.277 | 0.006 | 0.019 |
| 1.81 | 0.92 | 0.15 | 0.9 | 1.3 | 10 | 0.52 | 6 | 0.277 | 0.005 | -0.008 |
| 1.81 | 0.92 | 0.2 | 0.9 | 1.3 | 10 | 0.52 | 6 | 0.277 | 0.005 | -0.03 |
| 1.81 | 0.92 | 0.25 | 0.9 | 1.3 | 10 | 0.52 | 6 | 0.277 | 0.005 | -0.053 |
| 1.81 | 0.92 | 0.3 | 0.9 | 1.3 | 10 | 0.52 | 6 | 0.277 | 0.006 | -0.075 |
| 1.81 | 0.92 | 0.35 | 0.9 | 1.3 | 10 | 0.52 | 6 | 0.277 | 0.008 | -0.099 |
| 1.81 | 0.92 | 0.4 | 0.9 | 1.3 | 10 | 0.52 | 6 | 0.277 | 0.01 | -0.126 |
| 1.81 | 0.92 | 0.45 | 0.9 | 1.3 | 10 | 0.52 | 6 | 0.277 | 0.013 | -0.154 |
| 1.81 | 0.92 | 0.5 | 0.9 | 1.3 | 10 | 0.52 | 6 | 0.277 | 0.015 | -0.179 |
| 1.81 | 0.92 | 0.122 | 0.05 | 1.3 | 10 | 0.52 | 6 | 0.277 | 0.012 | 0.152 |
| 1.81 | 0.92 | 0.122 | 0.15 | 1.3 | 10 | 0.52 | 6 | 0.277 | 0.011 | 0.055 |
| 1.81 | 0.92 | 0.122 | 0.25 | 1.3 | 10 | 0.52 | 6 | 0.277 | 0.01 | 0.031 |
| 1.81 | 0.92 | 0.122 | 0.35 | 1.3 | 10 | 0.52 | 6 | 0.277 | 0.009 | 0.02 |
| 1.81 | 0.92 | 0.122 | 0.45 | 1.3 | 10 | 0.52 | 6 | 0.277 | 0.008 | 0.014 |
| 1.81 | 0.92 | 0.122 | 0.55 | 1.3 | 10 | 0.52 | 6 | 0.277 | 0.007 | 0.01 |
| 1.81 | 0.92 | 0.122 | 0.65 | 1.3 | 10 | 0.52 | 6 | 0.277 | 0.007 | 0.008 |
| 1.81 | 0.92 | 0.122 | 0.75 | 1.3 | 10 | 0.52 | 6 | 0.277 | 0.006 | 0.006 |
| 1.81 | 0.92 | 0.122 | 0.85 | 1.3 | 10 | 0.52 | 6 | 0.277 | 0.005 | 0.005 |
| 1.81 | 0.92 | 0.122 | 0.95 | 1.3 | 10 | 0.52 | 6 | 0.277 | 0.005 | 0.004 |
| 1.81 | 0.92 | 0.122 | 0.9 | 1.5 | 10 | 0.52 | 6 | 0.277 | 0.005 | 0.004 |
| 1.81 | 0.92 | 0.122 | 0.9 | 2.5 | 10 | 0.52 | 6 | 0.277 | 0.005 | 0.003 |
| 1.81 | 0.92 | 0.122 | 0.9 | 3.5 | 10 | 0.52 | 6 | 0.277 | 0.005 | 0.002 |
| 1.81 | 0.92 | 0.122 | 0.9 | 4.5 | 10 | 0.52 | 6 | 0.277 | 0.005 | 0.002 |
| 1.81 | 0.92 | 0.122 | 0.9 | 5.5 | 10 | 0.52 | 6 | 0.277 | 0.005 | 0.002 |
| 1.81 | 0.92 | 0.122 | 0.9 | 6.5 | 10 | 0.52 | 6 | 0.277 | 0.005 | 0.002 |
| 1.81 | 0.92 | 0.122 | 0.9 | 7.5 | 10 | 0.52 | 6 | 0.277 | 0.005 | 0.002 |
| 1.81 | 0.92 | 0.122 | 0.9 | 8.5 | 10 | 0.52 | 6 | 0.277 | 0.005 | 0.002 |
| 1.81 | 0.92 | 0.122 | 0.9 | 9.5 | 10 | 0.52 | 6 | 0.277 | 0.005 | 0.002 |
| 1.81 | 0.92 | 0.122 | 0.9 | 10.5 | 10 | 0.52 | 6 | 0.277 | 0.005 | 0.002 |
| 1.81 | 0.92 | 0.122 | 0.9 | 1.3 | 1.5 | 0.52 | 6 | 0.277 | 0.003 | 0.001 |
| 1.81 | 0.92 | 0.122 | 0.9 | 1.3 | 2.5 | 0.52 | 6 | 0.277 | 0.003 | 0.002 |
| 1.81 | 0.92 | 0.122 | 0.9 | 1.3 | 3.5 | 0.52 | 6 | 0.277 | 0.004 | 0.002 |
| 1.81 | 0.92 | 0.122 | 0.9 | 1.3 | 4.5 | 0.52 | 6 | 0.277 | 0.004 | 0.003 |
| 1.81 | 0.92 | 0.122 | 0.9 | 1.3 | 5.5 | 0.52 | 6 | 0.277 | 0.004 | 0.003 |
| 1.81 | 0.92 | 0.122 | 0.9 | 1.3 | 6.5 | 0.52 | 6 | 0.277 | 0.005 | 0.003 |
| 1.81 | 0.92 | 0.122 | 0.9 | 1.3 | 7.5 | 0.52 | 6 | 0.277 | 0.005 | 0.004 |
| 1.81 | 0.92 | 0.122 | 0.9 | 1.3 | 8.5 | 0.52 | 6 | 0.277 | 0.005 | 0.004 |
| 1.81 | 0.92 | 0.122 | 0.9 | 1.3 | 9.5 | 0.52 | 6 | 0.277 | 0.005 | 0.004 |
| 1.81 | 0.92 | 0.122 | 0.9 | 1.3 | 10.5 | 0.52 | 6 | 0.277 | 0.005 | 0.004 |
| 1.81 | 0.92 | 0.122 | 0.9 | 1.3 | 10 | 0.05 | 6 | 0.277 | -0.005 | -0.017 |
| 1.81 | 0.92 | 0.122 | 0.9 | 1.3 | 10 | 0.15 | 6 | 0.277 | 0.001 | -0.005 |
| 1.81 | 0.92 | 0.122 | 0.9 | 1.3 | 10 | 0.25 | 6 | 0.277 | 0.003 | -0.001 |
| 1.81 | 0.92 | 0.122 | 0.9 | 1.3 | 10 | 0.35 | 6 | 0.277 | 0.004 | 0.001 |
| 1.81 | 0.92 | 0.122 | 0.9 | 1.3 | 10 | 0.45 | 6 | 0.277 | 0.005 | 0.003 |
| 1.81 | 0.92 | 0.122 | 0.9 | 1.3 | 10 | 0.55 | 6 | 0.277 | 0.005 | 0.004 |
| 1.81 | 0.92 | 0.122 | 0.9 | 1.3 | 10 | 0.65 | 6 | 0.277 | 0.006 | 0.005 |
| 1.81 | 0.92 | 0.122 | 0.9 | 1.3 | 10 | 0.75 | 6 | 0.277 | 0.006 | 0.006 |
| 1.81 | 0.92 | 0.122 | 0.9 | 1.3 | 10 | 0.85 | 6 | 0.277 | 0.006 | 0.007 |
| 1.81 | 0.92 | 0.122 | 0.9 | 1.3 | 10 | 0.95 | 6 | 0.277 | 0.006 | 0.007 |
| 1.81 | 0.92 | 0.122 | 0.9 | 1.3 | 10 | 0.52 | 1.5 | 0.277 | 0.005 | 0.004 |
| 1.81 | 0.92 | 0.122 | 0.9 | 1.3 | 10 | 0.52 | 2.5 | 0.277 | 0.005 | 0.004 |
| 1.81 | 0.92 | 0.122 | 0.9 | 1.3 | 10 | 0.52 | 3.5 | 0.277 | 0.005 | 0.004 |
| 1.81 | 0.92 | 0.122 | 0.9 | 1.3 | 10 | 0.52 | 4.5 | 0.277 | 0.005 | 0.004 |
| 1.81 | 0.92 | 0.122 | 0.9 | 1.3 | 10 | 0.52 | 5.5 | 0.277 | 0.005 | 0.004 |
| 1.81 | 0.92 | 0.122 | 0.9 | 1.3 | 10 | 0.52 | 6.5 | 0.277 | 0.005 | 0.004 |
| 1.81 | 0.92 | 0.122 | 0.9 | 1.3 | 10 | 0.52 | 7.5 | 0.277 | 0.005 | 0.004 |
| 1.81 | 0.92 | 0.122 | 0.9 | 1.3 | 10 | 0.52 | 8.5 | 0.277 | 0.005 | 0.004 |
| 1.81 | 0.92 | 0.122 | 0.9 | 1.3 | 10 | 0.52 | 9.5 | 0.277 | 0.005 | 0.004 |
| 1.81 | 0.92 | 0.122 | 0.9 | 1.3 | 10 | 0.52 | 10.5 | 0.277 | 0.005 | 0.004 |
| 1.81 | 0.92 | 0.122 | 0.9 | 1.3 | 10 | 0.52 | 6 | 0.25 | 0.005 | 0.001 |
| 1.81 | 0.92 | 0.122 | 0.9 | 1.3 | 10 | 0.52 | 6 | 0.3 | 0.005 | 0.007 |
| 1.81 | 0.92 | 0.122 | 0.9 | 1.3 | 10 | 0.52 | 6 | 0.35 | 0.005 | 0.013 |
| 1.81 | 0.92 | 0.122 | 0.9 | 1.3 | 10 | 0.52 | 6 | 0.4 | 0.005 | 0.02 |
| 1.81 | 0.92 | 0.122 | 0.9 | 1.3 | 10 | 0.52 | 6 | 0.45 | 0.005 | 0.026 |
| 1.81 | 0.92 | 0.122 | 0.9 | 1.3 | 10 | 0.52 | 6 | 0.5 | 0.005 | 0.032 |
| 1.81 | 0.92 | 0.122 | 0.9 | 1.3 | 10 | 0.52 | 6 | 0.55 | 0.005 | 0.037 |
| 1.81 | 0.92 | 0.122 | 0.9 | 1.3 | 10 | 0.52 | 6 | 0.6 | 0.005 | 0.042 |
| 1.81 | 0.92 | 0.122 | 0.9 | 1.3 | 10 | 0.52 | 6 | 0.65 | 0.005 | 0.045 |
| 1.81 | 0.92 | 0.122 | 0.9 | 1.3 | 10 | 0.52 | 6 | 0.7 | 0.005 | 0.049 |

Bias formula = $\exp(\text{mean}(\log(\widehat{OR}))) - RR$ .
**Abbreviations:** *H*: health and healthcare-seeking behaviors; *I*: Infection status; *I* = 2: SARS-CoV-2 infection status; OR, odds ratio; RR, risk ratio; *T*: SARS-CoV-2 diagnostic tests; TND, test-negative design; *V*: COVID-19 vaccination status.
**Note:** Cells highlighted in yellow indicate the selected value for the parameter being varied.

**Table S5:** Bias in odds ratio (OR) estimates for **measured** symptomatic COVID-19 (*OR_COVID_*) and **measured** SARS-CoV-2 infection (*OR_infect_*) relative to their target parameters (*RR_COVID_* and *RR_infect_*, respectively) after individually varying the strength of two or three selected parameters (assuming symptomatic individuals are tested exclusively with NAATs and asymptomatic individuals are tested exclusively with antigen tests).

| OR HV | OR VT | I2 Prevalence | Bias Classical | Bias Alternative |
| --- | --- | --- | --- | --- |
| 1.5 | 0.95 | 0.122 | 0.006 | -0.004 |
| 2.5 | 0.85 | 0.122 | 0.003 | 0.022 |
| 3.5 | 0.75 | 0.122 | 0.002 | 0.050 |
| 4.5 | 0.65 | 0.122 | 0.001 | 0.083 |
| 5.5 | 0.55 | 0.122 | 0.000 | 0.120 |
| 6.5 | 0.45 | 0.122 | 0.000 | 0.166 |
| 7.5 | 0.35 | 0.122 | 0.000 | 0.225 |
| 8.5 | 0.25 | 0.122 | -0.001 | 0.307 |
| 9.5 | 0.15 | 0.122 | -0.001 | 0.439 |
| 10.5 | 0.05 | 0.122 | -0.001 | 0.734 |
| 1.5 | 0.92 | 0.5 | 0.018 | -0.186 |
| 2.5 | 0.92 | 0.45 | 0.009 | -0.140 |
| 3.5 | 0.92 | 0.4 | 0.003 | -0.093 |
| 4.5 | 0.92 | 0.35 | -0.001 | -0.047 |
| 5.5 | 0.92 | 0.3 | -0.003 | -0.004 |
| 6.5 | 0.92 | 0.25 | -0.003 | 0.037 |
| 7.5 | 0.92 | 0.2 | -0.003 | 0.079 |
| 8.5 | 0.92 | 0.15 | -0.002 | 0.122 |
| 9.5 | 0.92 | 0.1 | 0.001 | 0.175 |
| 10.5 | 0.92 | 0.05 | 0.007 | 0.255 |
| 1.81 | 0.95 | 0.5 | 0.015 | -0.181 |
| 1.81 | 0.85 | 0.45 | 0.013 | -0.149 |
| 1.81 | 0.75 | 0.4 | 0.010 | -0.113 |
| 1.81 | 0.65 | 0.35 | 0.008 | -0.078 |
| 1.81 | 0.55 | 0.3 | 0.006 | -0.044 |
| 1.81 | 0.45 | 0.25 | 0.005 | -0.011 |
| 1.81 | 0.35 | 0.2 | 0.005 | 0.025 |
| 1.81 | 0.25 | 0.15 | 0.005 | 0.068 |
| 1.81 | 0.15 | 0.1 | 0.006 | 0.131 |
| 1.81 | 0.05 | 0.05 | 0.010 | 0.265 |
| 1.5 | 0.95 | 0.5 | 0.018 | -0.188 |
| 2.5 | 0.85 | 0.45 | 0.009 | -0.134 |
| 3.5 | 0.75 | 0.4 | 0.003 | -0.076 |
| 4.5 | 0.65 | 0.35 | -0.001 | -0.017 |
| 5.5 | 0.55 | 0.3 | -0.003 | 0.044 |
| 6.5 | 0.45 | 0.25 | -0.003 | 0.111 |
| 7.5 | 0.35 | 0.2 | -0.003 | 0.191 |
| 8.5 | 0.25 | 0.15 | -0.002 | 0.295 |
| 9.5 | 0.15 | 0.1 | 0.001 | 0.452 |
| 10.5 | 0.05 | 0.05 | 0.007 | 0.766 |

Bias formula = $\exp(\text{mean}(\log(\widehat{OR}))) - RR$ .
**Abbreviations:** *H*: health and healthcare-seeking behaviors; *I*: Infection status; *I* = 2: SARS-CoV-2 infection status; OR, odds ratio; RR, risk ratio; *T*: SARS-CoV-2 diagnostic tests; TND, test-negative design; *V*: COVID-19 vaccination status.
**Note:** Cells highlighted in yellow indicate the selected value for the parameter being varied.

## Supplementary Figures

**Figure S1:**
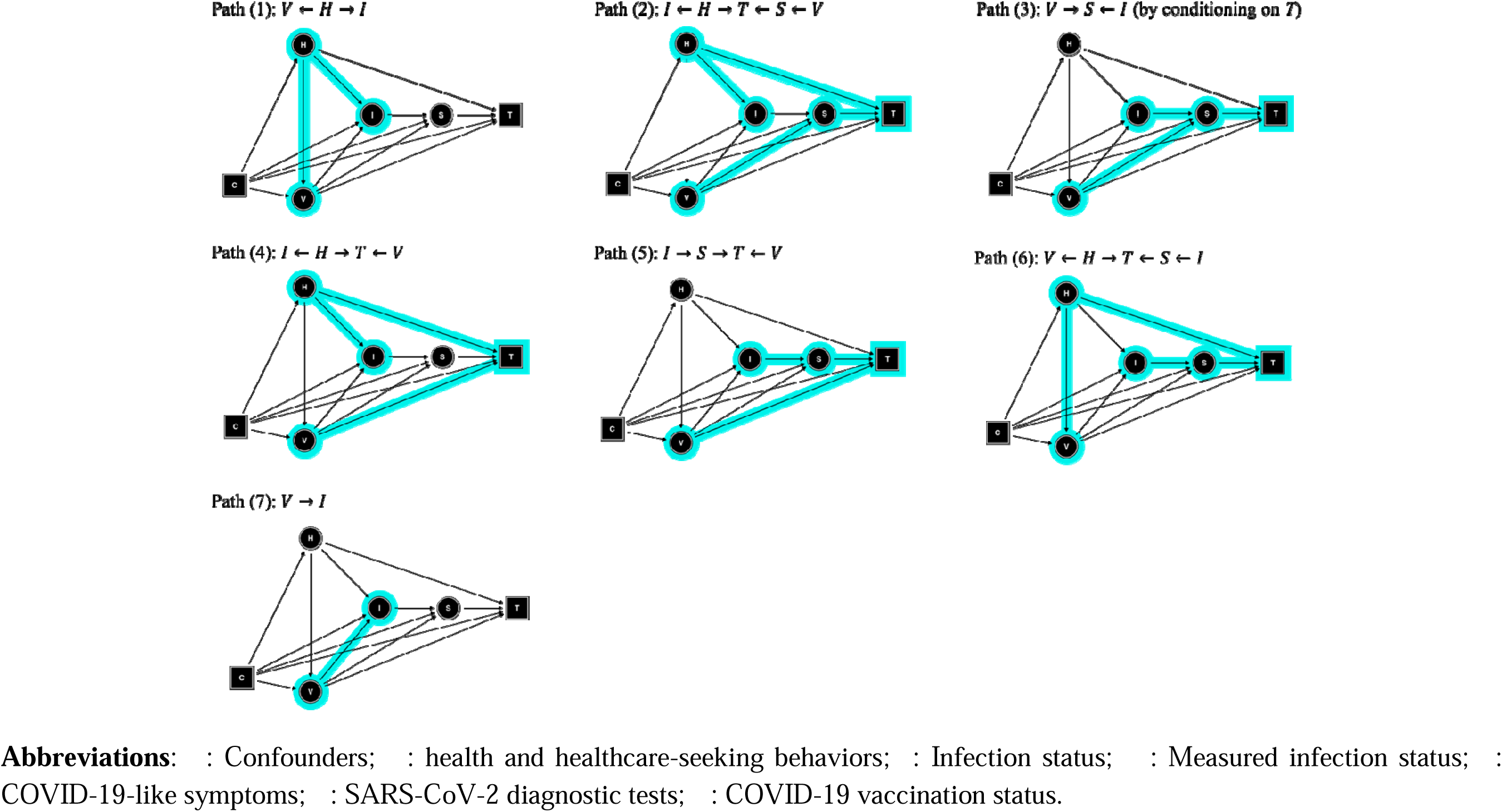
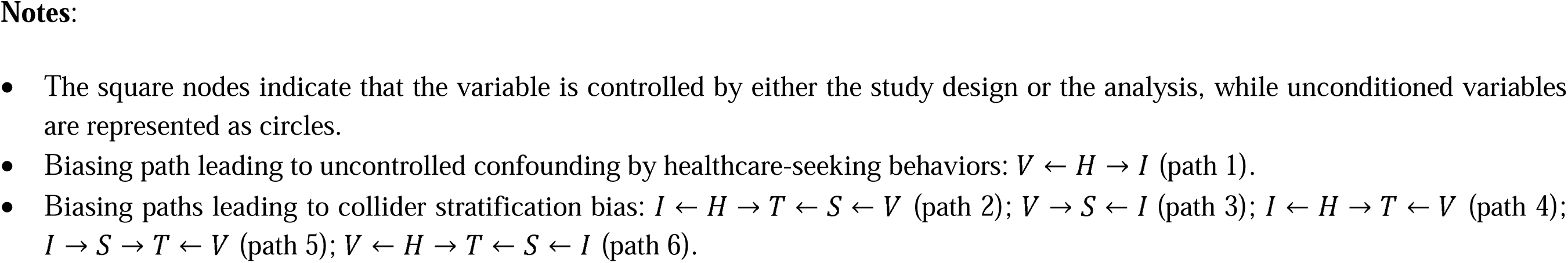
Causal directed acyclic graphs for “alternative” test-negative design studies of COVID-19 vaccine effectiveness – highlighting the remaining open paths between vaccination status and the outcome infection () after restricting the study sample to tested individuals ().

